# Drug allergy labels and complications after surgery: a prospective multi-centre cohort study

**DOI:** 10.64898/2026.06.04.26354882

**Authors:** Louise C Savic, Priya Dias, Jai Variale, Salma Begum, Kamran Khan, Alexander J Fowler, Vikas Kaura, Sarah-Louise Watson, Anna Littlejohns, Rupert M Pearse, Tom E F Abbott, the SAPPHIRE study investigators

## Abstract

**Background:** One in four surgical patients carries a drug allergy label, of which an estimated 90% are incorrect. Avoidance of first-choice drug therapies may lead to worse postoperative outcomes. We sought to determine the nature and extent of any association between drug allergy labels and postoperative complications.

**Methods:** A multicentre observational study in 21 NHS hospitals. Eligible patients were ≥18 years, undergoing common surgical procedures: primary hip or knee replacement; internal fixation of closed long bone fracture; colorectal resection; trans-urethral resection of prostate or bladder tumour; caesarean section; hysterectomy. Exclusion criteria: use of antibiotics in the two weeks prior to surgery, previous participation in the study. Primary outcome was postoperative complications within 30 days following surgery, a composite outcome comprising: all postoperative infections, anastomotic leak, acute respiratory distress syndrome, myocardial infarction, postoperative bleed, pulmonary embolism, stroke, antimicrobial side effects, death.

**Results:** Among 13,646 patients, 3924 (29%) carried ≥1 drug allergy labels. Labelled patients were more likely to develop postoperative complications (989/3924 (25%) vs 1926/9722 (20%); OR 1·21 [1·10-1·34]; p<0·001). They were more likely to develop surgical site infections (337/3924 (9%) vs 760/9722 (8%); OR 1·19 [1·03 -1·38]; p<0·018), and any postoperative infection (750/3924 (19%) vs 1472/9722 (15%); OR 1·24 [1·11-1·38] p<0·001). Labelled patients experienced increased risk of allergic drug reactions (31/3924 (0·01%) vs 29/9722 (<0·01%); OR 3·00 [1·77-5·09]; p<0·001), but no increase in mortality.

**Conclusions:** Drug allergy labels are common, but often incorrect. Labelled patients experience worse postoperative outcomes, including infective and non-infective complications and increased risk of allergic drug reactions.

**Trial registration:** Registered with ISRCTN registry, ISRCTN15775657.

## Introduction

One in four surgical patients carries a label of drug allergy, typically to antimicrobial and analgesic drugs.^(1)^ It is estimated that at least 90% of labels are not associated with a genuine drug allergy, but have been applied for other reasons such as minor side effects, symptoms of concomitant illness, unrelated non-allergic, idiosyncratic reactions, or for no clear, rational basis that can be identified. ^(1-6)^ Nevertheless, once a label has been recorded on the electronic health record this can be difficult to challenge even if demonstrably unlikely to be accurate. The label commits prescribers to using alternatives which may be less effective or have a more harmful side effect profile. The aim of this study was to determine whether there was an association between the presence of drug allergy labels and postoperative complications.

Previous work has demonstrated a link between penicillin allergy, the most prevalent label, and poor health outcomes. These effects are partly mediated through receipt of broad-spectrum antimicrobials which may be less effective at treating the underlying infection, and are associated with increased side effects including development of antimicrobial resistance. ^(7-11)^ A recent meta-analysis found that surgical patients with a label of beta-lactam allergy were at increased risk of surgical site infection.^(12)^ However, the relationship between other types of drug allergy label and complications among surgical patients remains uncertain.

We conducted a multicentre observational study using routinely collected data in a large, generalisable surgical population in the UK. We sought to determine the extent and nature of any association between the presence of drug allergy labels and the incidence of a wide range of postoperative complications.

## Methods

### Study design

We conducted a multi-centre observational cohort study to explore the association between the presence of drug allergy labels and incidence of surgical site infection and other surgical complications among patients undergoing one of six common surgical procedures (see below). This was a non-consenting study using routinely collected health data. The study was approved by the Health Research Authority and Health and Care Research Wales in February 2022 (REC reference 22/YH/0040, IRAS number 282161). The study protocol was registered with the ISRCTN registry, ISRCTN15775657.

### Participants

Eligible patients were aged 18 years or over, undergoing one of the following surgical procedures in participating secondary care hospitals: primary hip or knee replacement; internal fixation of a closed long bone fracture (upper or lower limb); colorectal resection; trans-urethral resection of prostate or bladder tumour; caesarean section; hysterectomy (vaginal or abdominal). These procedures were selected because they are performed frequently and therefore likely to provide generalisable findings. Open, robotic, laparoscopic, laparoscopically assisted and laparoscopic procedures converted to open, were all eligible versions of the above procedures. Exclusion criteria were the use of antibiotics in the two weeks prior to surgery and previous participation in the study. Follow-up data were collected for 30 days after the surgical procedure.

### Procedures

Age was recorded as a categorical variable in ten-year epochs from the age of 18 years. Sex was recorded as male or female. The presence of any drug allergy label was recorded, and the name(s) of the implicated drug(s). Smoking status was dichotomised as current or not current smoker. American Society of Anesthesiologists Physical Status (ASA) Classification was categorised as class one to four. We recorded the following chronic diseases: obesity (defined as BMI >30), liver cirrhosis, heart failure, hypertension, asthma or COPD, diabetes mellitus, active cancer, coronary artery disease, previous stroke, or peripheral vascular disease. We grouped surgical procedures into the six categories detailed above, and investigators recorded the use of open surgical technique, whether there was wound contamination, and the duration of surgery. We recorded the type of antimicrobial drugs administered as prophylaxis and to treat a confirmed or assumed infection.

### Exposure

The exposure of interest was the presence of a drug allergy label regardless of whether the diagnosis was correct and/or had been confirmed by an allergy specialist.

### Outcomes

The primary outcome was postoperative complications within 30 days following surgery, defined according to the US Centers for Disease Control criteria.^(13)^ This was a composite of all postoperative infections (superficial surgical site infection, deep surgical site infection, organ space infection, pneumonia, urinary tract infection, blood stream infection, intracranial infection, or infection with uncertain source), anastomotic leak, acute respiratory distress syndrome, myocardial infarction, postoperative bleed, pulmonary embolism, stroke, antimicrobial side effects (allergic reaction, acute kidney injury, diarrhoeal illness, hearing loss, tinnitus or vertigo), and death. Secondary outcomes were surgical site infections, any postoperative infection, number of antimicrobial drug types administered, antimicrobial side effects, allergic drug reactions, and death, within 30 days.

### Sample size calculation

The incidence of postoperative complications in exposed patients was expected to be 15%, and 10% in unexposed. Assuming a conservative estimate of prevalence of allergy labels of 10% a sample size of 13,720 patients would detect this difference with a power of 80% and significance level of 5% (two-sided test).

### Statistical analysis

The analysis was conducted in accordance with a pre-specified statistical analysis plan. We report a summary of descriptive data stratified by the presence or absence of a drug allergy label. This includes the clinical care received in terms of the surgical procedure category and mode of anaesthesia. We summarise the category and frequency of drug allergy labels. Continuous data are presented as mean (SD) or median (IQR) and categorical data as number (%). Hypothesis testing was not conducted on baseline data. Missing data were handled according to a pre-specified strategy, depending on the extent and nature of missing data. The primary analysis only included participants with complete data. If missing data were substantial and assumed to be missing at random (MAR), multiple imputation techniques were employed using the MICE R package (14).

To investigate associations between the presence of drug allergy labels and postoperative complications, we performed a mixed effects logistic regression model with a random intercept for site. Covariates were included in the model based on clinical plausibility using a forced simultaneous entry approach. These were: age, sex, surgical procedure, current smoker, diabetes mellitus, ASA, wound contamination, duration of surgery, active cancer, liver cirrhosis, obesity, and frailty. For the analysis of mortality, we included hypertension, heart failure, coronary artery disease, peripheral vascular disease, and asthma or COPD as covariates. A mixed effects logistic regression model was then fitted to estimate the effect of the drug allergy labels on surgical site infections, adjusted for any confounders.

For the secondary analyses, we repeated the above approach using the outcomes of surgical site infections, all post-operative infections, allergic reactions and death within 30 days after surgery. Lastly, we conducted pre-specified sensitivity analyses using the presence of any antimicrobial allergy, and specifically the presence of a penicillin allergy label, as the exposures of interest, with the outcomes of postoperative complications, surgical site infection, all postoperative infections, total number of antimicrobial agents administered, incidence of antimicrobial side effects, and (for antimicrobial allergy labels) mortality, all within 30 days after surgery. Analyses were conducted using R V4.4.3; R Core Team, 2024 (15). The results are reported with 95% confidence intervals (CI) and p-values. A significance level of p < 0.05 is considered statistically significant.

## Results

We included 13,720 eligible patients between March 2022 and October 2024. See Figure 1 for study flow diagram. Baseline and clinical care characteristics of the cohort, stratified by the presence of a drug allergy label, are described in Table 1.

**Table 1:**
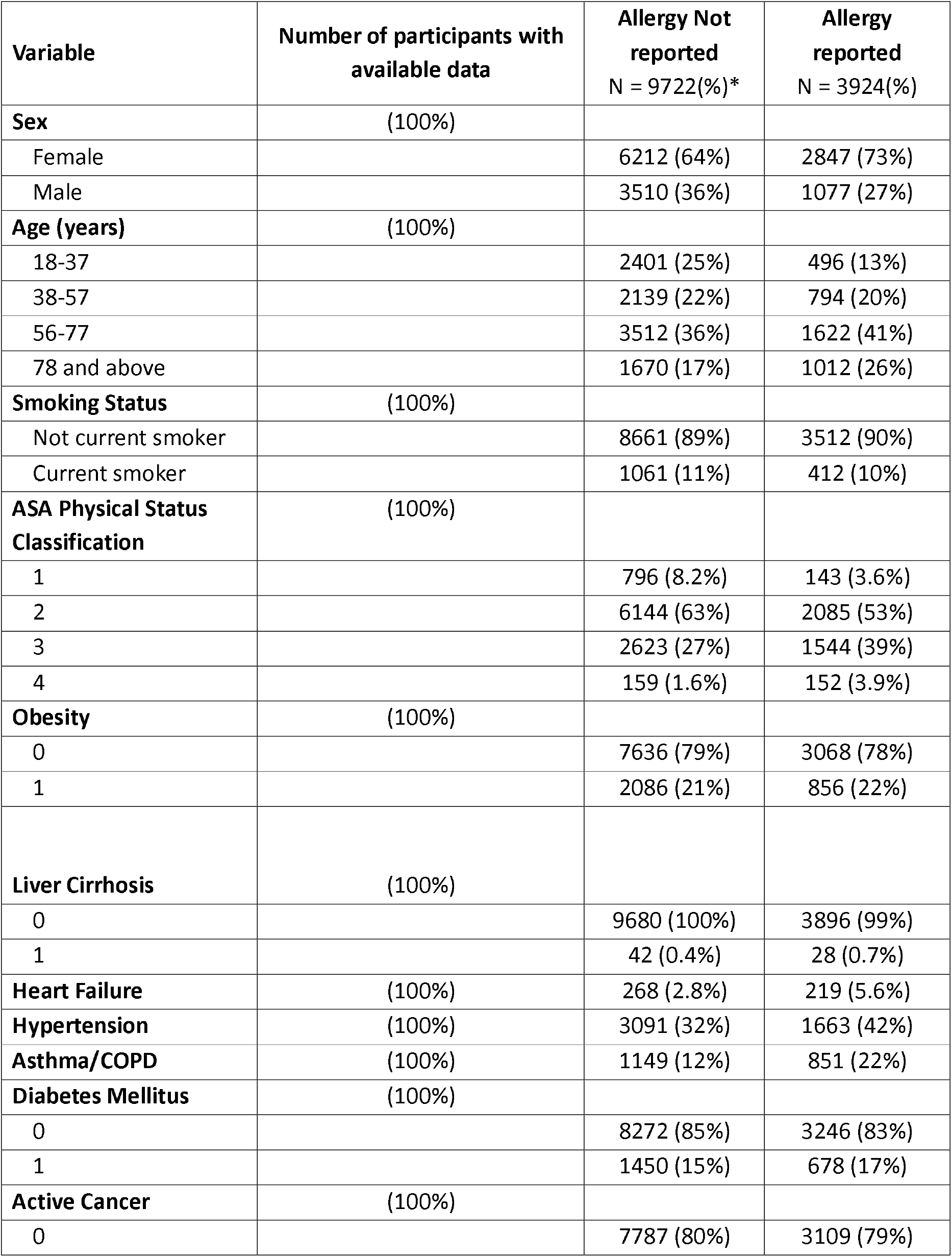
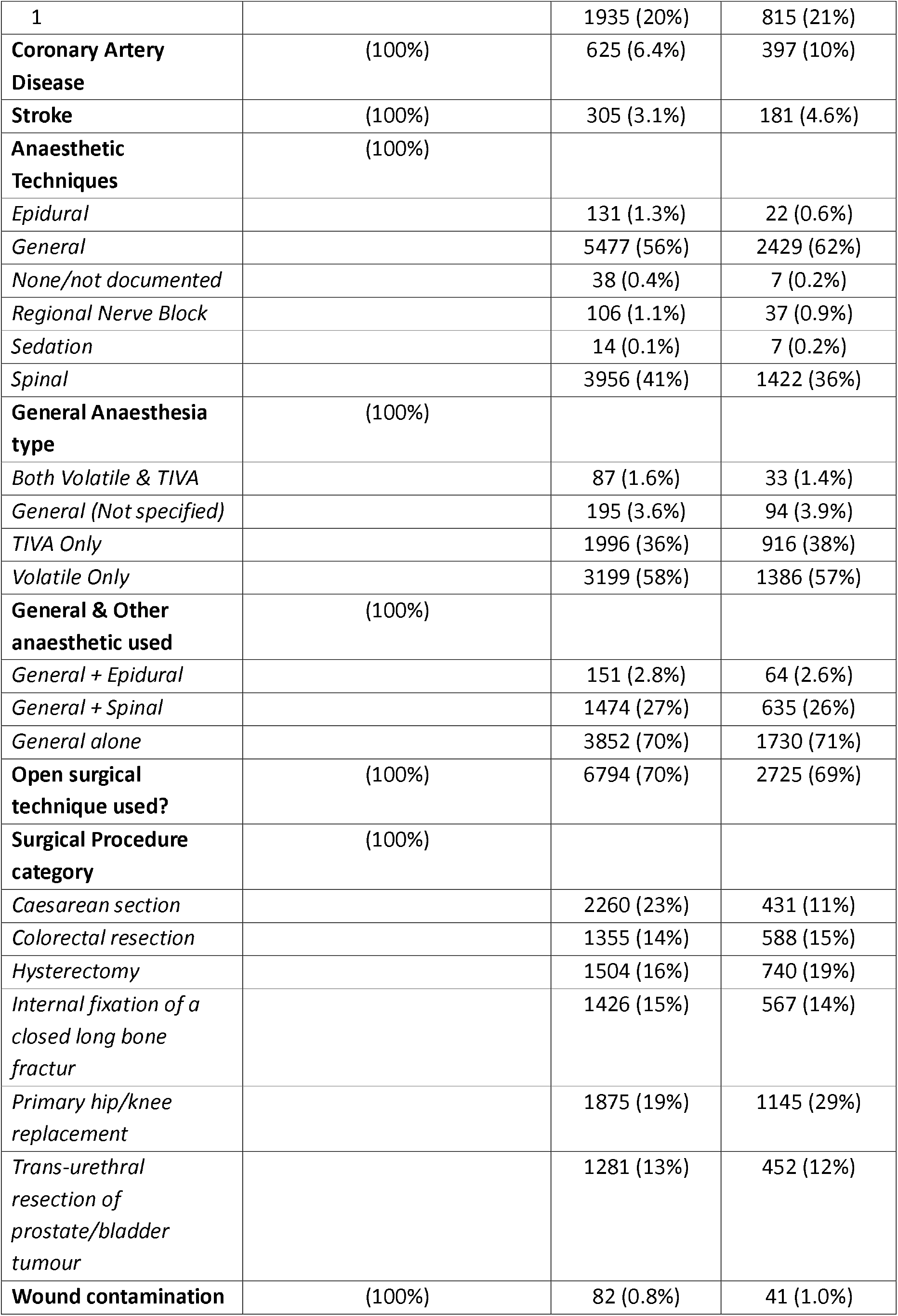
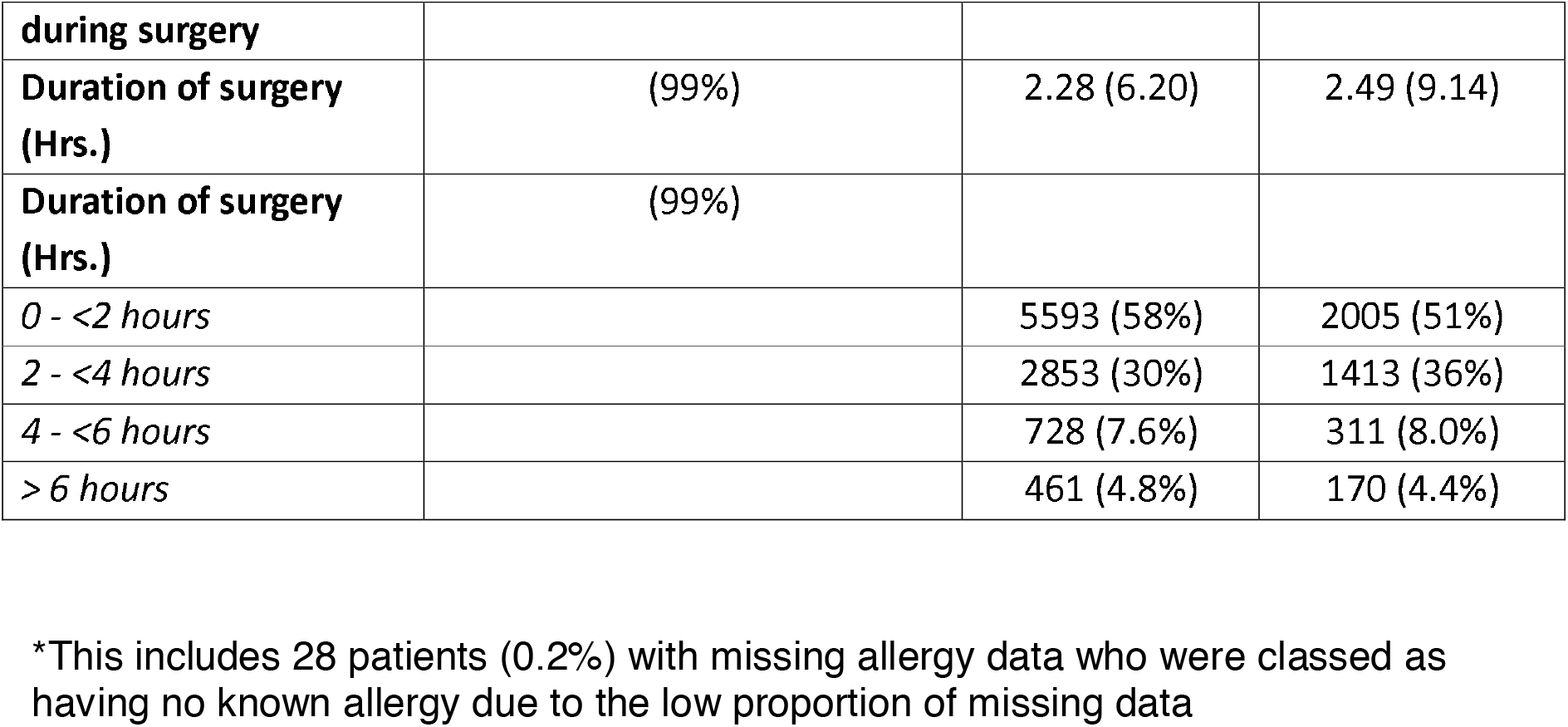
Baseline and clinical care characteristics of included patients, stratified by the presence of one or more drug allergy labels.

**Figure 1.**
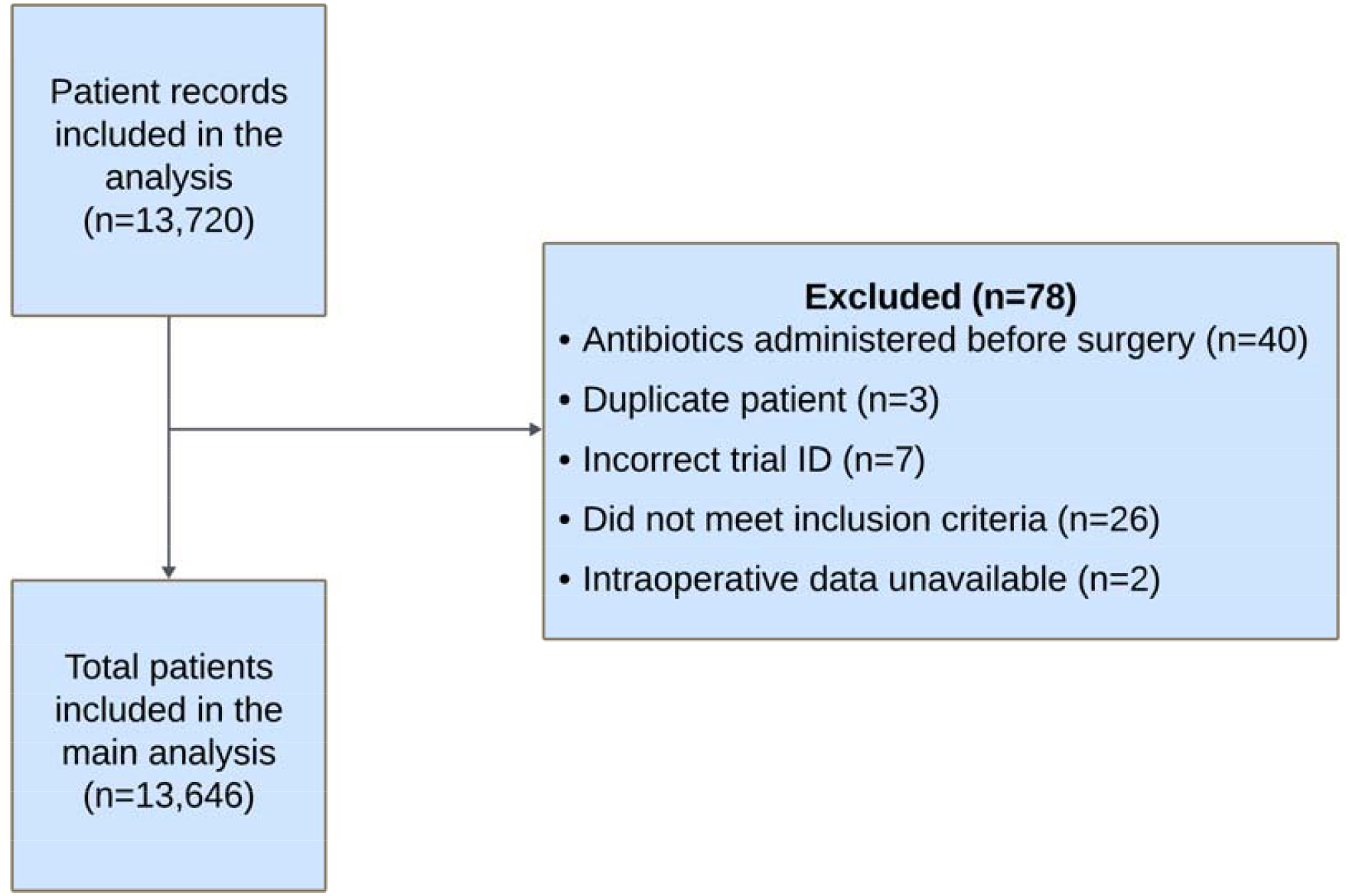
Flow diagram showing the number of patients included in the analysis.

**Figure 2.**
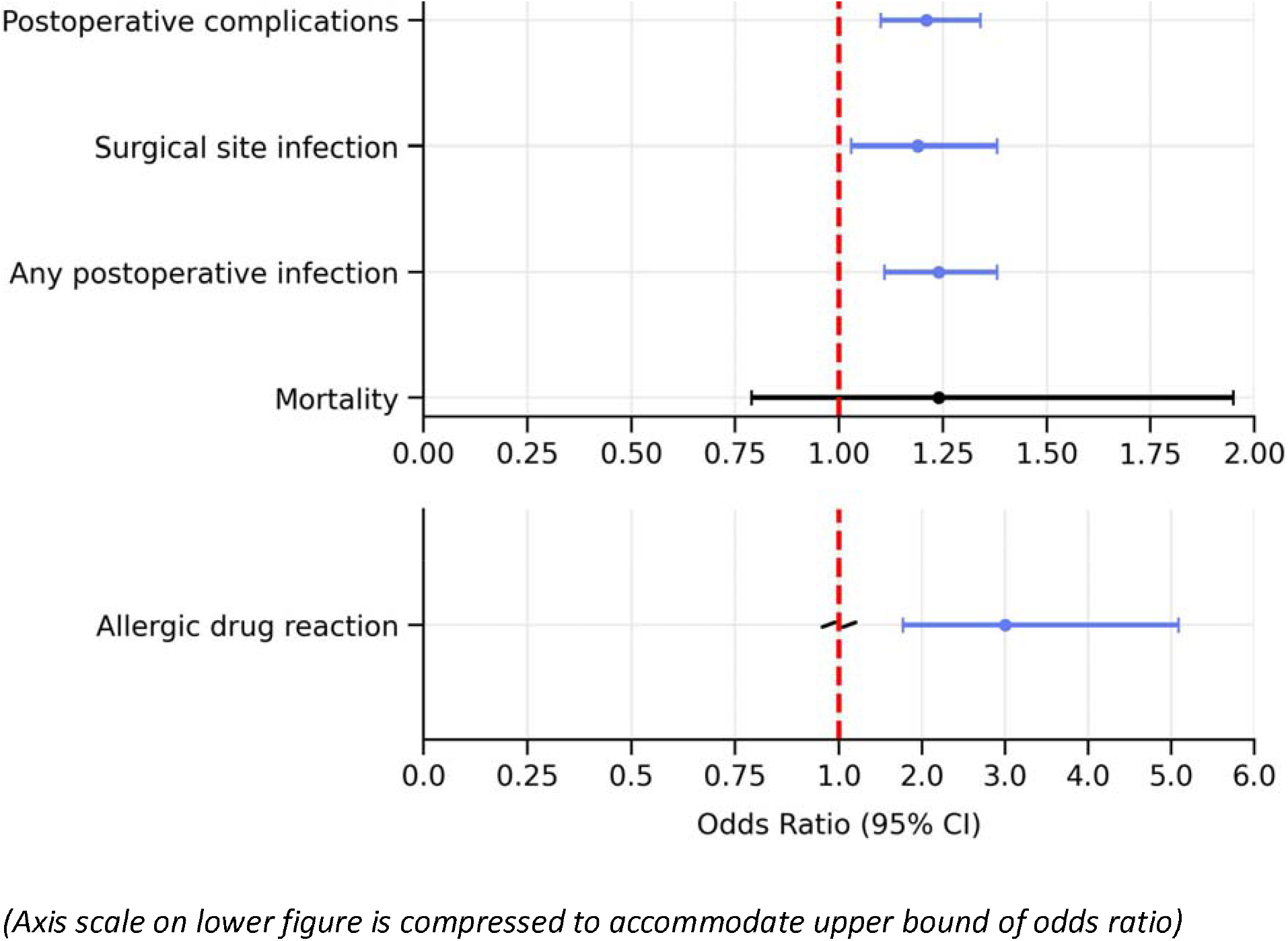
Forest plot summarising mixed effects logistic regression and negative binomial models used to estimate the effect of the drug allergy labels on surgical outcomes. Dependent variables are incidence of postoperative complications, surgical site infections, all infections, death, and allergic reactions, within 30 days after surgery. Significant findings are highlighted in blue. The x axis shows odds ratios and the error bars show 95% confidence intervals. Full multivariable models presented in Supplemental Files.

A total of 3924/13,720 (29%) of patients reported one or more drug allergy labels. The single most frequently reported allergy label was to penicillin (1573/3924, 40%). A total of 2169/3924 (55%) reported an antimicrobial allergy label, and 1190/3924 (30%) reported allergy to analgesics (including opioids and non-steroidal anti-inflammatory drugs, NSAIDs) (Table 2). A summary of the frequency of clinical outcomes, stratified by the presence of drug allergy labels is provided in Table 3 and a summary of primary and secondary analyses in Table 4.

**Table 2:** Categories and numbers of drug allergy labels among patients with one or more labels.

| Category of allergy | Number of patients reporting the allergy, expressed as % of patients with a drug allergy label (3924) and % of the entire patient cohort (13,646) |
| --- | --- |
| Any antimicrobial agent | 2169 (55%, **16%**) |
| Penicillin | 1573 (40%, **12%**) |
| Analgesics (including NSAIDs and opioids) | 1190 (30%, **9%**) |
| Other antimicrobial agents (non-penicillin) | 874 (22%, **6%**) |
| Adhesive tapes or plasters | 87 (2%, **<1%**) |
| Induction agents and neuromuscular blocking agents | 11 (<1%, **<1%**) |
| Drugs not commonly used during perioperative period* | 1344 (34%, **10%**) |
| Mis-recording of non-drug allergies** | 83 (2%, **1%**) |

% in bold type indicates % of total patient cohort of 13,646; (Total number of reported allergies (5171) is greater than number of patients reporting allergies (3924) due to reporting of multiple allergies); \*e.g. AdcalD3, ACEI, contrast media, intravenous iron preparations; \*\*e.g. aeroallergens, food allergens

**Table 3:** Summary of all postoperative complications within 30 days after surgery, stratified by the presence or absence of one or more drug allergy labels.

| Characteristic | Dug allergy not reported<br>N = 9722<sup>1</sup> | Drug allergy reported<br>N = 3924 |
| --- | --- | --- |
| **Post-operative infections, n (%)** | **1469 (15.1%)** | **747 (19.0%)** |
| Surgical site infection, n (%) | 760 (7.8%) | 337 (8.6%) |
| Pneumonia, n (%) | 251 (2.6%) | 192 (4.9%) |
| UTI (Non-TURP/TURBT), n (%) | 297 (3.1%) | 179 (4.6%) |
| Bloodstream, n (%) | 57 (0.6%) | 24 (0.6%) |
| Intracranial, n (%) | 1 (<0.1%) | 1 (<0.1%) |
| Infection, source uncertain, n (%) | 278 (2.9%) | 113 (2.9%) |
| **Surgical site infections (SSI), n (%)** | **760 (7.8%)** | **337 (8.6%)** |
| Superficial, n (%) | 396 (4.1%) | 179 (4.6%) |
| Deep Surgical, n (%) | 137 (1.4%) | 67 (1.7%) |
| Organ Space, n (%) | 118 (1.2%) | 48 (1.2%) |
| UTI (TURP/TURBT), n (%) | 157 (1.6%) | 70 (1.8%) |
| **Post operative complications, n (%)** | **1900 (19.5%)** | **978 (24.9%)** |
| Deaths, n (%) | 54 (0.6%) | 36 (0.9%) |
| Post-operative infections, n (%) | 1469 (15.1%) | 747 (19.0%) |
| Allergic reaction, n (%) | 29 (0.3%) | 31 (0.8%) |
| Acute kidney injury, n (%) | 528 (5.4%) | 292 (7.4%) |
| Diarrhoea, n (%) | 46 (0.5%) | 18 (0.5%) |
| Hearing loss, n (%) | 4 (<0.1%) | 2 (<0.1%) |
| Tinnitus, n (%) | 1 (<0.1%) | 1 (<0.1%) |
| Vertigo, n (%) | 4 (<0.1%) | 4 (0.1%) |
| Anastomotic leak, n (%) | 41 (0.4%) | 19 (0.5%) |
| ARDS, n (%) | 8 (<0.1%) | 4 (0.1%) |
| Myocardial Infarction, n (%) | 11 (0.1%) | 3 (<0.1%) |
| Postoperative bleed, n (%) | 97 (1.0%) | 40 (1.0%) |
| Pulmonary embolism, n (%) | 24 (0.2%) | 17 (0.4%) |
| Stroke, n (%) | 10 (0.1%) | 12 (0.3%) |
| Side effects, n (%) | 564 (5.8%) | 311 (7.9%) |
| **Side effects of antimicrobials, n (%)** | **564 (5.8%)** | **311 (7.9%)** |
| Acute kidney injury, n (%) | 528 (5.4%) | 292 (7.4%) |
| Diarrhoea, n (%) | 46 (0.5%) | 18 (0.5%) |
| Hearing loss, n (%) | 4 (<0.1%) | 2 (<0.1%) |
| Tinnitus, n (%) | 1 (<0.1%) | 1 (<0.1%) |
| Vertigo, n (%) | 4 (<0.1%) | 4 (0.1%) |

<sup>1</sup>n (%)

**Table 4:** Summary of primary and secondary outcomes, stratified by the presence of drug allergy labels.

| Outcomes | Drug allergy reported<sup>a</sup> | Drug allergy *not* | Effect size in patients with | P-value |
| --- | --- | --- | --- | --- |

|  |  | reported<sup>b</sup> | reported drug<br>allergy<br>Odds ratio (95% CI) |  |
| --- | --- | --- | --- | --- |
| **Primary outcome** |  |  |  |  |
| Postoperative complications<sup>c</sup><br>n (%) | 989 (25%) | 1926 (20%) | 1.21 (1.10 – 1.34) | **<0.001<sup>3</sup>** |
| **Secondary outcomes** |  |  |  |  |
| Surgical site infection | 337 (8.6%) | 760 (7.8%) | 1.19 (1.03 – 1.38) | **<0.018<sup>1</sup>** |
| Postoperative infections<sup>d</sup><br>n (%) | 750 (19%) | 1472 (15%) | 1.24 (1.11 – 1.38) | **<0.001<sup>2</sup>** |
| Allergic reactions<br>n (%) | 31 (0.8%) | 29 (0.3%) | 3.00 (1.77 – 5.09) | **<0.001** |
| Death<br>n (%) | 36 (0.9%) | 54 (0.6%) | 1.24 (0.79 – 1.95) | 0.35 |

**1-4** See Supplementary Files S1-4 for full regression models for each analysis
**a,b** Data available for 100% of patients
**c** Composite outcome of postoperative infection (surgical site infection, pneumonia, urinary tract infection, abscess or collection, bloodstream infection, intracranial infection, meningitis, infection of uncertain source), anastomotic leak, ARDS, myocardial infarction, postoperative bleed, pulmonary embolism, stroke, diarrhoeal illness, hearing loss, vertigo, tinnitus, acute kidney injury, allergic reaction, death
**d** Postoperative infection: composite outcome of surgical site infection, pneumonia, urinary tract infection, abscess or collection, bloodstream infection, intracranial infection, meningitis, infection of uncertain source

Among patients with a drug allergy label, 989/3924 (25%) developed a postoperative complication within 30 days after surgery, compared with 1926/9722 (20%) of those without a drug allergy label (OR 1.21 [1.10-1.34]; p<0.001) (Table 4 and Supplementary table S1). Surgical site infections were more common among patients with a drug allergy label compared to those without a label (Allergy label 337/3924 (8·6%) patients vs No allergy label 760/9722 (7·8%) patients; OR 1.19 [1.03-1.38]; p<0.018). The incidence of any postoperative infection was more common in those with a drug allergy label (Allergy label 750/3924 (19%) patients vs No label 1472/9722 (15%) patients; OR 1.24 [1.11-1.38]; p<0.001), and there was an increased risk of allergic reaction in those with a label (Allergy label 31/3924 (0.8%) vs No label 29/9722 (0.3%); OR 3.00 [1.77-5.09]; p<0.001). The incidence of postoperative mortality was similar between groups (Table 4 and Supplementary Files, Tables S2-5).

In the sensitivity analyses, patients with any antibiotic allergy label experienced an increased risk of postoperative complications compared to those without (OR 1.19 [1.06 – 1.34]; p<0.004. Those with an antibiotic allergy also experienced greater risk of surgical site infections (OR 1.22 [1.02 – 1.45]; p<0.027) and of all postoperative infections (OR 1.27 [1.12-1.45]; p<0.001). Patients with an antibiotic allergy label were administered a greater number of different antimicrobial agents but did not experience an increased rate of antimicrobial side effects. There was no difference in mortality at 30 days between the two groups. Patients with a penicillin allergy label experienced an increased risk of postoperative complications (OR 1.25 [1.09 – 1.43]; p=0.002), surgical site infections (OR 1.25 [1.02 – 1.53]; p<0.030), and all postoperative infections (OR 1.30 [1.12 – 1.51]; p<0.001). They were administered a greater total number of different antimicrobial agents but did not experience an increase in antimicrobial side effects. See Supplementary Files, Tables S6-16.

## Discussion

The principal finding of this study is that one in four surgical patients carries a drug allergy label, and this group experiences an increased risk of complications within 30 days after surgery. There is also an increased risk of postoperative infection, including surgical site infection, and greater likelihood of perioperative allergic drug reactions. This is the first time such associations have been defined for a large, generalisable surgical population. Whilst the effect sizes are modest, the very large number of patients reporting drug allergy creates a substantial burden of associated harm for health systems. Since the overwhelming majority of drug labels do not reflect true allergy, much of this harm is avoidable.

Drug allergy labels are a global public health issue.^(16)^ An estimated 36% of patients carry a label, and surgical patients are more likely to report drug allergy than other groups. ^(1, 17, 18)^ At least 90% of labels are found to be incorrect when tested. ^(1, 4, 19, 20)^ In line with global estimates, we found penicillin allergy to be the most prevalent label, with analgesics and other antimicrobials allergy comprising the remainder. ^(9, 21, 22)^ The presence of beta lactam allergy labels has been associated with increased infections, increased risk of antimicrobial resistance, longer hospital stays, increased rates of treatment failure and readmission, and higher all-cause mortality.(7-10, 23-26) A recent systematic review and meta-analysis demonstrated increased risk of surgical site infection (OR 1.60 95%CI 1.27-2.01 p<0.0001) in patients with a beta lactam allergy label. ^(12)^ However, the included studies were mostly single centre, examining small numbers of patients undergoing single surgical procedures, with no adjustment for important covariates known to be associated with negative health outcomes The impact of other drug allergy labels on surgical outcomes has not been systematically investigated until now. ^(1, 27, 28).^ Our data describe a clear association between the presence of any drug allergy label and the risk of postoperative complications including infections, myocardial infarction, stroke, pulmonary embolus and risk of perioperative allergic reactions, in a generalisable surgical population. Causal mechanisms for the observed associations cannot be determined from our data. Plausible pathways include increased risk of infection and development of resistant organism secondary to poor antimicrobial stewardship, or ineffective postoperative pain relief due to analgesic allergy labels, leading to poor cough strength, reduced mobility and poor wound healing. These in turn can directly contribute to the risk of chest infections, thrombo-embolic events and other complications. Further research is needed to elucidate these pathways and determine whether pre-operative removal of drug allergy labels modifies these poor outcomes.

Our study has several strengths. The surgical procedures selected, large number of included sites, and high data completion rates, means results are likely to be generalisable to surgical patients in high income countries. Our reported estimates of the number and categories of drug allergy labels are consistent with previous epidemiological work, providing face validity for these data. By adjusting for the presence of confounding variables known to be important for postoperative complications, we were able to provide more accurate estimates for these effects. However, as for all observational studies, we were unable to adjust for unmeasured confounders. In addition, the lack of primary care data for many patients means that complications occurring post-discharge and managed in the community, were incompletely captured in our data. This may have under-estimated the incidence of postoperative complications, resulting in a smaller estimate of the degree of association between variables. Lastly, the international generalisability of some of our data may be limited by varying patterns of antimicrobial use.

## Conclusion

Regardless of accuracy of the allergy diagnosis, patients with drug allergy labels experience worse outcomes after surgery, with an increase in postoperative complications. Rather than reducing the risk of allergic reactions to drugs, the presence of a label is associated with an increased risk of this. Research is urgently needed to understand the negative impact of drug allergy labels on patient care.

## Supporting information

Investigator List

## Data Availability

All data produced in the present study are available upon reasonable request to the authors

## Authors’ contributions

LS and TA conceived the study. LS, PD, AF, VK, RP, TA were responsible for study design. LS, TA, PD, SB, JV, SW, AL and the SAPPHIRE study group were responsible for data collection. KK was responsible for data analysis. LS, TA and RP were responsible for data interpretation. LS and TA wrote the first draft of the manuscript. All authors revised the manuscript for important intellectual content and approved the final version. KK had full access to the data and act as guarantor.

## Declaration of interests

AJF holds a National Institute for Health Research Doctoral Research fellowship (DRF-2018-11-ST2-062. RP is an NIHR senior investigator. TA is supported by an NIHR DSE (NIHR305701); has received research funding from NIHR (CL-2021-19-501), Barts Charity, the Academy of Medical Sciences, The Royal College of Anaesthetists and British Journal of Anaesthesia; has received honoraria from MSD, Edwards Life Sciences and Elsevier; and is Social Media Editor of the British Journal of Anaesthesia. All other authors report no financial relationships with any organisations that might have an interest in the submitted work in the previous three years, no other relationships or activities that could appear to have influenced the submitted work.

## Transparency declaration

LS/TA affirm that the manuscript is an honest, accurate, and transparent account of the study being reported; that no important aspects of the study have been omitted; and that any discrepancies from the study as planned have been explained.

## Funding

National Institute for Health and Care Research (NIHR) Clinical Lectureship (CL-2021-19-501), NIHR Doctoral Research Fellowship (DRF-2020-301454) and National Institute for Academic Anaesthesia (WKRO-2020-0020). The funding sources had no role in the study design, data collection, analysis, interpretation, or writing the report.

## Notes

### Competing Interest Statement

The authors have declared no competing interest.

### Author Declarations

The study was approved by the Health Research Authority and Health and Care Research Wales in February 2022 (Research Ethics Committee reference 22/YH/0040, IRAS number 282161).

