## Supplementary Files 1-16 for "Drug allergy labels and complications after surgery: a prospective multi-centre cohort study"

| **S1** | Mixed-effects logistic regression examining the association between drug allergy labels and all postoperative complications within 30 days after surgery |
| --- | --- |
| **S2** | Mixed-effects logistic regression examining the association between drug allergy labels and surgical site infections within 30 days after surgery |
| **S3** | Mixed-effects logistic regression examining the association between drug allergy labels and all postoperative infections within 30 days after surgery |
| **S4** | Mixed-effects logistic regression examining the association between drug allergy labels and mortality within 30 days of surgery |
| **S5** | Mixed-effects logistic regression examining the association between drug allergy labels and allergic reactions within 30 days after surgery |
| **S6** | Mixed-effects logistic regression examining the association between antimicrobial allergy labels and all postoperative complications within 30 days of surgery |
| **S7** | Mixed-effects logistic regression examining the association between antimicrobial allergy labels and surgical site infections within 30 days of surgery |
| **S8** | Mixed-effects negative binomial regression examining the association between antimicrobial allergy labels and all postoperative infections within 30 days after surgery |
| **S9** | Mixed-effects binomial regression examining the association between antimicrobial allergy labels and number of antimicrobial agents administered from recruitment until 30 days after surgery |
| **S10** | Mixed effects logistic regression examining the association between antimicrobial allergy labels and incidence of antimicrobial side effects within 30 days after surgery |
| **S11** | Mixed effects logistic regression examining the association between antimicrobial allergy labels and mortality within 30 days after surgery |
| **S12** | Mixed-effects logistic regression examining the association between penicillin allergy labels and all postoperative complications within 30 days of surgery |
| **S13** | Mixed-effects logistic regression examining the association between penicillin allergy labels and surgical site infections within 30 days of surgery |
| **S14** | Mixed-effects negative binomial regression examining the association between penicillin allergy labels and all postoperative infections within 30 days after surgery |
| **S15** | Mixed-effects binomial regression examining the association between penicillin allergy labels and number of antimicrobial agents administered from recruitment until 30 days after surgery |
| **S16** | Mixed effects logistic regression examining the association between penicillin allergy labels and incidence of antimicrobial side effects within 30 days after surgery |

**S1:** **Mixed-effects logistic regression examining the association between drug allergy labels and postoperative complications within 30 days after surgery, adjusting for clinically important covariates**

| **Characteristic**^1^ | **OR** **(95% CI)** | **p-value** |
| --- | --- | --- |
| **(Intercept)** | 0.05 (0.03 to 0.07) | **<0.001** |
| **Drug allergy** |  |  |
| Absent | — |  |
| Present | 1.21 (1.10 to 1.34) | **<0.001** |
| **Age** |  |  |
| 18-37 | — |  |
| 38-57 | 1.13 (0.94 to 1.35) | 0.20 |
| 56-77 | 1.26 (1.03 to 1.54) | **0.025** |
| 78 and above | 1.81 (1.45 to 2.27) | **<0.001** |
| **Sex** |  |  |
| Female | — |  |
| Male | 1.18 (1.05 to 1.33) | **0.006** |
| **Current smoker** |  |  |
| Current Smoker(N) | — |  |
| Current Smoker(Y) | 1.19 (1.03 to 1.37) | **0.016** |
| **Surgical procedure** |  |  |
| Primary hip/knee replacement | — |  |
| Caesarean section | 1.97 (1.55 to 2.50) | **<0.001** |
| Colorectal resection | 1.82 (1.51 to 2.20) | **<0.001** |
| Hysterectomy | 1.37 (1.14 to 1.64) | **<0.001** |
| Internal fixation of a closed long bone fracture | 1.01 (0.85 to 1.19) | 0.93 |
| Trans-urethral resection of prostate/bladder tumour | 1.11 (0.92 to 1.34) | 0.26 |
| **Wound contamination** |  |  |
| No | — |  |
| Yes | 2.30 (1.55 to 3.43) | **<0.001** |
| **Surgery time (hrs)** |  |  |
| 0 - <2 hours | — |  |
| 2 - <4 hours | 1.40 (1.23 to 1.59) | **<0.001** |
| 4 - <6 hours | 1.75 (1.43 to 2.13) | **<0.001** |
| > 6 hours | 3.13 (2.48 to 3.95) | **<0.001** |
| **Frailty** |  |  |
| Non-frail | — |  |
| Mild Frailty | 1.14 (0.99 to 1.31) | 0.062 |
| Moderate Frailty | 1.40 (1.18 to 1.66) | **<0.001** |
| Severe Frailty | 3.29 (2.33 to 4.65) | **<0.001** |
| **ASA Physical Status Classification** |  |  |
| 1 | — |  |
| 2 | 1.31 (1.05 to 1.65) | **0.019** |
| 3 | 1.90 (1.49 to 2.44) | **<0.001** |
| 4 | 3.43 (2.41 to 4.87) | **<0.001** |
| **Diabetes Mellitus** | 1.24 (1.11 to 1.40) | **<0.001** |
| **Heart Failure** | 1.18 (0.95 to 1.47) | 0.14 |
| **Stroke** | 0.98 (0.79 to 1.23) | 0.89 |
| **Hypertension** | 1.08 (0.97 to 1.20) | 0.14 |
| **Coronary Artery Disease** | 1.05 (0.89 to 1.23) | 0.58 |
| **Asthma/COPD** | 1.19 (1.05 to 1.34) | **0.006** |
| **Peripheral Vascular Disease** | 0.98 (0.73 to 1.30) | 0.88 |
| **Active Cancer** | 0.97 (0.86 to 1.10) | 0.68 |
| **Liver Cirrhosis** | 0.96 (0.54 to 1.69) | 0.89 |
| **Obesity** | 1.17 (1.05 to 1.31) | **0.005** |
| **centreid.sd__(Intercept)** | 0.51 (NA to NA) |  |

^1^Random Effects: σ² (Residual) = 3.29, τ00 (centreid) = 0.26, ICC = 0.07, N (centreid) = 23, Observations = 13,458, Marginal R²/Conditional R² = 0.100/0.165

Abbreviations: CI = Confidence Interval, OR = Odds Ratio

**S2: Mixed-effects logistic regression examining the association between drug allergy labels and surgical site infections within 30 days of surgery, adjusting for clinically important covariates.**

| **Characteristic**^1^ | **OR** **(95% CI)** | **p-value** |
| --- | --- | --- |
| **(Intercept)** | 0.01 (0.01 to 0.02) | **<0.001** |
| **Drug allergy** |  |  |
| Absent | — |  |
| Present | 1.19 (1.03 to 1.38) | **0.018** |
| **Age** |  |  |
| 18-37 | — |  |
| 38-57 | 0.86 (0.67 to 1.10) | 0.24 |
| 56-77 | 0.79 (0.60 to 1.04) | 0.10 |
| 78 and above | 0.77 (0.56 to 1.07) | 0.12 |
| **Sex** |  |  |
| Female | — |  |
| Male | 1.29 (1.07 to 1.56) | **0.007** |
| **Current smoker** |  |  |
| Current Smoker(N) | — |  |
| Current Smoker(Y) | 1.15 (0.94 to 1.41) | 0.17 |
| **Surgical procedure** |  |  |
| Primary hip/knee replacement | — |  |
| Caesarean section | 3.55 (2.42 to 5.19) | **<0.001** |
| Colorectal resection | 3.71 (2.71 to 5.08) | **<0.001** |
| Hysterectomy | 2.51 (1.84 to 3.43) | **<0.001** |
| Internal fixation of a closed long bone fracture | 0.87 (0.61 to 1.23) | 0.44 |
| Trans-urethral resection of prostate/bladder tumour | 6.26 (4.60 to 8.52) | **<0.001** |
| **Wound contamination** |  |  |
| No | — |  |
| Yes | 2.17 (1.33 to 3.55) | **0.002** |
| **Surgery time (hrs)** |  |  |
| 0 - <2 hours | — |  |
| 2 - <4 hours | 1.58 (1.26 to 1.97) | **<0.001** |
| 4 - <6 hours | 1.99 (1.48 to 2.69) | **<0.001** |
| > 6 hours | 2.98 (2.16 to 4.13) | **<0.001** |
| **Frailty** |  |  |
| Non-frail | — |  |
| Mild Frailty | 0.82 (0.66 to 1.02) | 0.069 |
| Moderate Frailty | 0.68 (0.50 to 0.93) | **0.016** |
| Severe Frailty | 1.45 (0.78 to 2.70) | 0.24 |
| **ASA Physical Status Classification** |  |  |
| 1 | — |  |
| 2 | 1.32 (0.95 to 1.84) | 0.10 |
| 3 | 1.89 (1.32 to 2.71) | **<0.001** |
| 4 | 2.73 (1.54 to 4.84) | **<0.001** |
| **Diabetes Mellitus** | 1.24 (1.05 to 1.47) | **0.012** |
| **Heart Failure** | 1.08 (0.75 to 1.57) | 0.67 |
| **Stroke** | 0.86 (0.59 to 1.26) | 0.44 |
| **Hypertension** | 0.89 (0.76 to 1.04) | 0.13 |
| **Coronary Artery Disease** | 0.99 (0.77 to 1.27) | 0.91 |
| **Asthma/COPD** | 1.06 (0.88 to 1.27) | 0.56 |
| **Peripheral Vascular Disease** | 0.70 (0.42 to 1.16) | 0.17 |
| **Active Cancer** | 0.84 (0.72 to 1.00) | **0.047** |
| **Liver Cirrhosis** | 1.00 (0.44 to 2.27) | >0.99 |
| **Obesity** | 1.44 (1.23 to 1.68) | **<0.001** |
| **centreid.sd__(Intercept)** | 0.54 (NA to NA) |  |
| ^1^Random Effects: σ² (Residual) = 3.29, τ00 (centreid) = 0.29, ICC = 0.08, N (centreid) = 23, Observations = 13,458, Marginal R²/Conditional R² = 0.160/0.228 | | |
| Abbreviations: CI = Confidence Interval, OR = Odds Ratio | | |

**S3: Mixed-effects logistic regression examining the association between drug allergy labels and postoperative infections, adjusting for clinically important covariates.**

| **Characteristic**^1^ | **OR** **(95% CI)** | **p-value** |
| --- | --- | --- |
| **(Intercept)** | 0.03 (0.02 to 0.05) | **<0.001** |
| **drug_allergy** |  |  |
| Absent | — |  |
| Present | 1.24 (1.11 to 1.38) | **<0.001** |
| **Age** |  |  |
| 18-37 | — |  |
| 38-57 | 0.99 (0.81 to 1.21) | 0.93 |
| 56-77 | 1.12 (0.90 to 1.39) | 0.32 |
| 78 and above | 1.42 (1.12 to 1.82) | **0.004** |
| **Sex** |  |  |
| Female | — |  |
| Male | 1.15 (1.01 to 1.31) | **0.036** |
| **current_smoker** |  |  |
| Current Smoker(N) | — |  |
| Current Smoker(Y) | 1.31 (1.13 to 1.52) | **<0.001** |
| **Surgical procedure** |  |  |
| Primary hip/knee replacement | — |  |
| Caesarean section | 2.25 (1.73 to 2.93) | **<0.001** |
| Colorectal resection | 2.30 (1.86 to 2.83) | **<0.001** |
| Hysterectomy | 1.80 (1.48 to 2.21) | **<0.001** |
| Internal fixation of a closed long bone fracture | 1.01 (0.84 to 1.22) | 0.91 |
| Trans-urethral resection of prostate/bladder tumour | 1.47 (1.19 to 1.82) | **<0.001** |
| **woundcontamination_operation** |  |  |
| No | — |  |
| Yes | 2.10 (1.38 to 3.19) | **<0.001** |
| **Surgery time (hrs)** |  |  |
| 0 - <2 hours | — |  |
| 2 - <4 hours | 1.39 (1.20 to 1.60) | **<0.001** |
| 4 - <6 hours | 1.76 (1.42 to 2.17) | **<0.001** |
| > 6 hours | 2.63 (2.05 to 3.37) | **<0.001** |
| **Frailty** |  |  |
| Non-frail | — |  |
| Mild Frailty | 1.12 (0.96 to 1.31) | 0.14 |
| Moderate Frailty | 1.33 (1.10 to 1.61) | **0.004** |
| Severe Frailty | 2.96 (2.07 to 4.24) | **<0.001** |
| **ASA Physical Status Classification** |  |  |
| 1 | — |  |
| 2 | 1.27 (0.99 to 1.64) | 0.060 |
| 3 | 1.78 (1.36 to 2.34) | **<0.001** |
| 4 | 3.34 (2.29 to 4.86) | **<0.001** |
| **Diabetes Mellitus** | 1.15 (1.01 to 1.31) | **0.029** |
| **Heart Failure** | 1.03 (0.81 to 1.32) | 0.81 |
| **Stroke** | 1.06 (0.83 to 1.35) | 0.62 |
| **Hypertension** | 0.99 (0.88 to 1.11) | 0.90 |
| **Coronary Artery Disease** | 1.00 (0.84 to 1.20) | 0.99 |
| **Asthma/COPD** | 1.24 (1.09 to 1.41) | **0.001** |
| **Peripheral Vascular Disease** | 1.04 (0.76 to 1.42) | 0.81 |
| **Active Cancer** | 0.92 (0.81 to 1.05) | 0.22 |
| **Liver Cirrhosis** | 0.60 (0.30 to 1.21) | 0.15 |
| **Obesity** | 1.26 (1.12 to 1.42) | **<0.001** |
| **centreid.sd__(Intercept)** | 0.62 (NA to NA) |  |
| ^1^Random Effects: σ² (Residual) = 3.29, τ00 (centreid) = 0.39, ICC = 0.11, N (centreid) = 23, Observations = 13,447, Marginal R²/Conditional R² = 0.083/0.180 | | |

**S4:** **Mixed-effects logistic regression examining the association between drug allergy labels and mortality within 30 days of surgery, adjusting for clinically important covariates.**

| **Characteristic**^1^ | **OR** **(95% CI)** | **p-value** |
| --- | --- | --- |
| **(Intercept)** | 0.00 (0.00 to 0.00) | **<0.001** |
| **drug_allergy** |  |  |
| Absent | — |  |
| Present | 1.24 (0.79 to 1.95) | 0.35 |
| **Sex** |  |  |
| Female | — |  |
| Male | 2.27 (1.44 to 3.59) | **<0.001** |
| **current_smoker** |  |  |
| Current Smoker(N) | — |  |
| Current Smoker(Y) | 0.49 (0.19 to 1.22) | 0.13 |
| **woundcontamination_operation** |  |  |
| No | — |  |
| Yes | 1.48 (0.20 to 11.1) | 0.70 |
| **Frailty** |  |  |
| Non-frail | — |  |
| Mild Frailty | 3.48 (1.73 to 6.99) | **<0.001** |
| Moderate Frailty | 13.8 (7.54 to 25.2) | **<0.001** |
| Severe Frailty | 53.9 (26.4 to 110) | **<0.001** |
| **Diabetes Mellitus** | 0.87 (0.50 to 1.49) | 0.60 |
| **Heart Failure** | 0.68 (0.33 to 1.43) | 0.31 |
| **Stroke** | 0.88 (0.42 to 1.84) | 0.74 |
| **Hypertension** | 1.00 (0.63 to 1.58) | >0.99 |
| **Coronary Artery Disease** | 1.50 (0.87 to 2.57) | 0.14 |
| **Asthma/COPD** | 1.34 (0.81 to 2.21) | 0.26 |
| **Peripheral Vascular Disease** | 1.74 (0.79 to 3.85) | 0.17 |
| **Active Cancer** | 1.60 (1.00 to 2.55) | 0.051 |
| **Liver Cirrhosis** | 1.03 (0.13 to 8.19) | 0.98 |
| **Obesity** | 1.09 (0.61 to 1.98) | 0.77 |
| **centreid.sd__(Intercept)** | 0.35 (NA to NA) |  |

^1^Random Effects: σ² (Residual) = 3.29, τ00 (centreid) = 0.12, ICC = 0.04, N (centreid) = 23, Observations = 13,538, Marginal R²/Conditional R² = 0.262/0.289

**S5: Mixed-effects logistic regression examining association between drug allergy labels and allergic reactions, adjusting for clinically important covariates.**

| **Characteristic**^1^ | **OR** **(95% CI)** | **p-value** |
| --- | --- | --- |
| **(Intercept)** | 0.00 (0.00 to 0.00) | **<0.001** |
| **drug_allergy** |  |  |
| Absent | — |  |
| Present | 3.00 (1.77 to 5.09) | **<0.001** |
| **Sex** |  |  |
| Female | — |  |
| Male | 1.08 (0.60 to 1.93) | 0.80 |
| **current_smoker** |  |  |
| Current Smoker(N) | — |  |
| Current Smoker(Y) | 0.87 (0.37 to 2.05) | 0.75 |
| **Frailty** |  |  |
| Non-frail | — |  |
| Mild Frailty | 0.64 (0.24 to 1.66) | 0.36 |
| Moderate Frailty | 1.94 (0.84 to 4.51) | 0.12 |
| Severe Frailty | 1.64 (0.21 to 12.7) | 0.64 |
| **Diabetes Mellitus** | 0.64 (0.30 to 1.39) | 0.26 |
| **Heart Failure** | 1.02 (0.29 to 3.60) | 0.97 |
| **Hypertension** | 0.81 (0.45 to 1.45) | 0.48 |
| **Coronary Artery Disease** | 1.92 (0.83 to 4.48) | 0.13 |
| **Asthma/COPD** | 0.82 (0.40 to 1.72) | 0.60 |
| **Stroke** | 0.81 (0.19 to 3.43) | 0.77 |
| **Peripheral Vascular Disease** | 0.67 (0.09 to 4.97) | 0.70 |
| **Active Cancer** | 2.11 (1.21 to 3.67) | **0.008** |
| **Obesity** | 1.13 (0.61 to 2.06) | 0.70 |
| **centreid.sd__(Intercept)** | 1.06 (NA to NA) |  |

^1^Random Effects: σ² (Residual) = 3.29, τ00 (centreid) = 1.13, ICC = 0.26, N (centreid) = 23, Observations = 13,585, Marginal R²/Conditional R² = 0.094/0.325

**S6 Mixed-effects logistic regression examining the association between antimicrobial allergy labels and all postoperative complications within 30 days of surgery**

| **Characteristic**^1^ | **OR** **(95% CI)** | **p-value** |
| --- | --- | --- |
| **(Intercept)** | 0.05 (0.03 to 0.07) | **<0.001** |
| **Antimicrobial label** |  |  |
| No antimicrobial label | — |  |
| Any Antimicrobial | 1.19 (1.06 to 1.34) | **0.004** |
| **Age** |  |  |
| 18-37 | — |  |
| 38-57 | 1.13 (0.94 to 1.35) | 0.20 |
| 56-77 | 1.26 (1.03 to 1.54) | **0.026** |
| 78 and above | 1.82 (1.45 to 2.28) | **<0.001** |
| **Sex** |  |  |
| Female | — |  |
| Male | 1.17 (1.04 to 1.31) | **0.010** |
| **Current smoker** |  |  |
| Current Smoker(N) | — |  |
| Current Smoker(Y) | 1.19 (1.03 to 1.37) | **0.019** |
| **Surgical procedure** |  |  |
| Primary hip/knee replacement | — |  |
| Caesarean section | 1.93 (1.52 to 2.45) | **<0.001** |
| Colorectal resection | 1.82 (1.51 to 2.20) | **<0.001** |
| Hysterectomy | 1.36 (1.14 to 1.63) | **<0.001** |
| Internal fixation of a closed long bone fracture | 1.00 (0.85 to 1.18) | 0.98 |
| Trans-urethral resection of prostate/bladder tumour | 1.11 (0.92 to 1.34) | 0.27 |
| **Wound contamination** |  |  |
| No | — |  |
| Yes | 2.29 (1.54 to 3.42) | **<0.001** |
| **Surgery time (hrs)** |  |  |
| 0 - <2 hours | — |  |
| 2 - <4 hours | 1.41 (1.24 to 1.60) | **<0.001** |
| 4 - <6 hours | 1.75 (1.44 to 2.14) | **<0.001** |
| > 6 hours | 3.14 (2.49 to 3.95) | **<0.001** |
| **Frailty** |  |  |
| Non-frail | — |  |
| Mild Frailty | 1.15 (1.00 to 1.32) | 0.053 |
| Moderate Frailty | 1.40 (1.18 to 1.66) | **<0.001** |
| Severe Frailty | 3.31 (2.34 to 4.67) | **<0.001** |
| **ASA Physical Status Classification** |  |  |
| 1 | — |  |
| 2 | 1.32 (1.05 to 1.67) | **0.016** |
| 3 | 1.93 (1.50 to 2.46) | **<0.001** |
| 4 | 3.48 (2.45 to 4.94) | **<0.001** |
| **Diabetes Mellitus** | 1.25 (1.11 to 1.40) | **<0.001** |
| **Heart Failure** | 1.19 (0.96 to 1.49) | 0.11 |
| **Stroke** | 0.99 (0.79 to 1.23) | 0.91 |
| **Hypertension** | 1.08 (0.98 to 1.20) | 0.13 |
| **Coronary Artery Disease** | 1.05 (0.90 to 1.24) | 0.52 |
| **Asthma/COPD** | 1.20 (1.06 to 1.35) | **0.003** |
| **Peripheral Vascular Disease** | 0.98 (0.74 to 1.31) | 0.89 |
| **Active Cancer** | 0.97 (0.86 to 1.10) | 0.64 |
| **Liver Cirrhosis** | 0.98 (0.55 to 1.72) | 0.93 |
| **Obesity** | 1.17 (1.05 to 1.31) | **0.005** |
| **centreid.sd__(Intercept)** | 0.50 (NA to NA) |  |

^1^Random Effects: σ² (Residual) = 3.29, τ00 (centreid) = 0.25, ICC = 0.07, N (centreid) = 23, Observations = 13,458, Marginal R²/Conditional R² = 0.099/0.164

**S7 Mixed-effects logistic regression examining the association between antimicrobial allergy labels and surgical site infections within 30 days of surgery**

| **Characteristic**^1^ | **OR** **(95% CI)** | **p-value** |
| --- | --- | --- |
| **(Intercept)** | 0.01 (0.01 to 0.02) | **<0.001** |
| **Antimicrobial label** |  |  |
| No antimicrobial label | — |  |
| Any Antimicrobial | 1.22 (1.02 to 1.45) | **0.027** |
| **Age** |  |  |
| 18-37 | — |  |
| 38-57 | 0.86 (0.67 to 1.10) | 0.23 |
| 56-77 | 0.79 (0.60 to 1.04) | 0.092 |
| 78 and above | 0.77 (0.56 to 1.07) | 0.12 |
| **Sex** |  |  |
| Female | — |  |
| Male | 1.28 (1.07 to 1.54) | **0.008** |
| **Current Smoker** |  |  |
| Current Smoker(N) | — |  |
| Current Smoker(Y) | 1.15 (0.94 to 1.41) | 0.18 |
| **Surgical procedure** |  |  |
| Primary hip/knee replacement | — |  |
| Caesarean section | 3.48 (2.38 to 5.08) | **<0.001** |
| Colorectal resection | 3.72 (2.72 to 5.08) | **<0.001** |
| Hysterectomy | 2.50 (1.84 to 3.41) | **<0.001** |
| Internal fixation of a closed long bone fracture | 0.86 (0.61 to 1.22) | 0.41 |
| Trans-urethral resection of prostate/bladder tumour | 6.25 (4.60 to 8.49) | **<0.001** |
| **Wound Contamination** |  |  |
| No | — |  |
| Yes | 2.17 (1.33 to 3.54) | **0.002** |
| **Surgery time (hrs)** |  |  |
| 0 - <2 hours | — |  |
| 2 - <4 hours | 1.58 (1.27 to 1.97) | **<0.001** |
| 4 - <6 hours | 1.99 (1.48 to 2.69) | **<0.001** |
| > 6 hours | 2.99 (2.16 to 4.14) | **<0.001** |
| **Frailty** |  |  |
| Non-frail | — |  |
| Mild Frailty | 0.82 (0.66 to 1.02) | 0.072 |
| Moderate Frailty | 0.68 (0.50 to 0.93) | **0.016** |
| Severe Frailty | 1.46 (0.78 to 2.72) | 0.23 |
| **ASA Physical Status Classification** |  |  |
| 1 | — |  |
| 2 | 1.32 (0.95 to 1.84) | 0.10 |
| 3 | 1.90 (1.33 to 2.72) | **<0.001** |
| 4 | 2.77 (1.56 to 4.89) | **<0.001** |
| **Diabetes Mellitus** | 1.25 (1.05 to 1.48) | **0.011** |
| **Heart Failure** | 1.09 (0.76 to 1.58) | 0.64 |
| **Stroke** | 0.86 (0.59 to 1.25) | 0.44 |
| **Hypertension** | 0.89 (0.76 to 1.04) | 0.14 |
| **Coronary Artery Disease** | 0.99 (0.77 to 1.27) | 0.94 |
| **Asthma/COPD** | 1.07 (0.89 to 1.28) | 0.49 |
| **Peripheral Vascular Disease** | 0.70 (0.42 to 1.17) | 0.17 |
| **Active Cancer** | 0.84 (0.71 to 0.99) | **0.042** |
| **Liver Cirrhosis** | 1.02 (0.45 to 2.32) | 0.97 |
| **Obesity** | 1.44 (1.23 to 1.68) | **<0.001** |
| **centreid.sd__(Intercept)** | 0.54 (NA to NA) |  |

^1^Random Effects: σ² (Residual) = 3.29, τ00 (centreid) = 0.29, ICC = 0.08, N (centreid) = 23, Observations = 13,458, Marginal R²/Conditional R² = 0.160/0.228

Abbreviations: CI = Confidence Interval, OR = Odds Ratio

**S8 Mixed-effects negative binomial regression examining the association between antimicrobial allergy labels and all postoperative infections within 30 days after surgery**

| **Characteristic**^1^ | **OR** **(95% CI)** | **p-value** |
| --- | --- | --- |
| **(Intercept)** | 0.04 (0.02 to 0.05) | **<0.001** |
| **Antimicrobial label** |  |  |
| No antimicrobial label | — |  |
| Any Antimicrobial | 1.27 (1.12 to 1.45) | **<0.001** |
| **Age** |  |  |
| 18-37 | — |  |
| 38-57 | 0.99 (0.81 to 1.20) | 0.90 |
| 56-77 | 1.11 (0.90 to 1.38) | 0.34 |
| 78 and above | 1.43 (1.12 to 1.82) | **0.004** |
| **Sex** |  |  |
| Female | — |  |
| Male | 1.14 (1.00 to 1.30) | **0.046** |
| **current smoker** |  |  |
| Current Smoker(N) | — |  |
| Current Smoker(Y) | 1.31 (1.12 to 1.52) | **<0.001** |
| **Surgical procedure** |  |  |
| Primary hip/knee replacement | — |  |
| Caesarean section | 2.22 (1.71 to 2.89) | **<0.001** |
| Colorectal resection | 2.30 (1.87 to 2.83) | **<0.001** |
| Hysterectomy | 1.80 (1.47 to 2.20) | **<0.001** |
| Internal fixation of a closed long bone fracture | 1.00 (0.83 to 1.21) | 0.98 |
| Trans-urethral resection of prostate/bladder tumour | 1.46 (1.19 to 1.81) | **<0.001** |
| **wound contamination** |  |  |
| No | — |  |
| Yes | 2.08 (1.37 to 3.17) | **<0.001** |
| **Surgery time (hrs)** |  |  |
| 0 - <2 hours | — |  |
| 2 - <4 hours | 1.39 (1.21 to 1.60) | **<0.001** |
| 4 - <6 hours | 1.76 (1.42 to 2.18) | **<0.001** |
| > 6 hours | 2.64 (2.06 to 3.38) | **<0.001** |
| **Frailty** |  |  |
| Non-frail | — |  |
| Mild Frailty | 1.13 (0.97 to 1.32) | 0.12 |
| Moderate Frailty | 1.33 (1.10 to 1.61) | **0.003** |
| Severe Frailty | 2.98 (2.08 to 4.27) | **<0.001** |
| **ASA Physical Status Classification** |  |  |
| 1 | — |  |
| 2 | 1.26 (0.98 to 1.62) | 0.068 |
| 3 | 1.78 (1.36 to 2.33) | **<0.001** |
| 4 | 3.34 (2.29 to 4.86) | **<0.001** |
| **Diabetes Mellitus** | 1.16 (1.02 to 1.32) | **0.027** |
| **Heart Failure** | 1.04 (0.82 to 1.34) | 0.74 |
| **Stroke** | 1.07 (0.84 to 1.36) | 0.60 |
| **Hypertension** | 1.00 (0.89 to 1.12) | 0.95 |
| **Coronary Artery Disease** | 1.01 (0.84 to 1.20) | 0.93 |
| **Asthma/COPD** | 1.25 (1.10 to 1.43) | **<0.001** |
| **Peripheral Vascular Disease** | 1.04 (0.76 to 1.42) | 0.81 |
| **Active Cancer** | 0.92 (0.81 to 1.05) | 0.21 |
| **Liver Cirrhosis** | 0.61 (0.30 to 1.24) | 0.17 |
| **Obesity** | 1.26 (1.12 to 1.42) | **<0.001** |
| **centreid.sd__(Intercept)** | 0.62 (NA to NA) |  |
| **Characteristic**^1^ | **OR** **(95% CI)** | **p-value** |
| **(Intercept)** | 0.04 (0.02 to 0.05) | **<0.001** |
| **Antimicrobial label** |  |  |
| No antimicrobial label | — |  |
| Any Antimicrobial | 1.27 (1.12 to 1.45) | **<0.001** |
| **Age** |  |  |
| 18-37 | — |  |
| 38-57 | 0.99 (0.81 to 1.20) | 0.90 |
| 56-77 | 1.11 (0.90 to 1.38) | 0.34 |
| 78 and above | 1.43 (1.12 to 1.82) | **0.004** |
| ^1^Random Effects: σ² (Residual) = 3.29, τ00 (centreid) = 0.38, ICC = 0.10, N (centreid) = 23, Observations = 13,458, Marginal R²/Conditional R² = 0.083/0.179 | | |

**S9 Mixed-effects binomial regression examining the association between antimicrobial allergy labels and number of antimicrobial agents administered from recruitment until 30 days after surgery**

| **Group** | **Characteristic**^1^ | **exp(Beta)** **(95% CI)** | **p-value** |
| --- | --- | --- | --- |
| **cond** | (Intercept) | 1.35 (1.23 to 1.48) | **<0.001** |
|  | Antimicrobial label |  |  |
|  | No antimicrobial label | — |  |
|  | Any Antimicrobial | 1.13 (1.09 to 1.16) | **<0.001** |
|  | Age |  |  |
|  | Age38-57 | 0.97 (0.93 to 1.02) | 0.26 |
|  | Age56-77 | 1.00 (0.95 to 1.06) | 0.98 |
|  | Age78 and above | 1.07 (1.00 to 1.13) | **0.045** |
|  | Sex |  |  |
|  | Male | 1.04 (1.01 to 1.07) | **0.021** |
|  | Current Smoker |  |  |
|  | Current Smoker(Y) | 1.04 (1.0 to 1.08) | 0.089 |
|  | Surgical procedure |  |  |
|  | Primary hip/knee replacement | — |  |
|  | Caesarean section | 1.26 (1.18 to 1.35) | **<0.001** |
|  | Colorectal resection | 1.50 (1.43 to 1.59) | **<0.001** |
|  | Hysterectomy | 1.12 (1.07 to 1.18) | **<0.001** |
|  | Internal fixation of a closed long bone fracture | 1.11 (1.06 to 1.17) | **<0.001** |
|  | Trans-urethral resection of prostate/bladder tumour | 0.88 (0.83 to 0.93) | **<0.001** |
|  | Wound Contamination |  |  |
|  | wound contamination | 1.09 (0.98 to 1.22) | 0.12 |
|  | Surgery time (hrs) |  |  |
|  | 0 - <2 hours | — |  |
|  | 2 - <4 hours | 1.07 (1.03 to 1.11) | **<0.001** |
|  | 4 - <6 hours | 1.13 (1.07 to 1.20) | **<0.001** |
|  | > 6 hours | 1.35 (1.26 to 1.44) | **<0.001** |
|  | Frailty |  |  |
|  | Non-frail | — |  |
|  | Mild Frailty | 1.01 (0.97 to 1.05) | 0.65 |
|  | Moderate Frailty | 1.05 (1.00 to 1.11) | **0.046** |
|  | Severe Frailty | 1.23 (1.11 to 1.37) | **<0.001** |
|  | ASA Physical Status Classification |  |  |
|  | 1 | — |  |
|  | 2 | 1.07 (1.01 to 1.13) | **0.016** |
|  | 3 | 1.14 (1.07 to 1.21) | **<0.001** |
|  | 4 | 1.31 (1.19 to 1.45) | **<0.001** |
|  | Diabetes Mellitus | 1.03 (0.99 to 1.06) | 0.12 |
|  | Heart Failure | 1.00 (0.93 to 1.07) | 0.97 |
|  | Stroke | 1.04 (0.97 to 1.11) | 0.29 |
|  | Hypertension | 0.98 (0.95 to 1.01) | 0.29 |
|  | Coronary Artery Disease | 1.01 (0.96 to 1.06) | 0.74 |
|  | Asthma/COPD | 1.06 (1.02 to 1.09) | **0.002** |
|  | Peripheral Vascular Disease | 0.99 (0.91 to 1.08) | 0.83 |
|  | Active Cancer | 0.98 (0.95 to 1.02) | 0.27 |
|  | Liver Cirrhosis | 0.96 (0.81 to 1.14) | 0.63 |
|  | Obesity | 1.03 (1.00 to 1.06) | 0.086 |
|  | centreid.sd__(Intercept) | 0.12 (NA to NA) |  |

^1^Random Effects: σ² (Residual) = 0.43, τ00 (centreid) = 0.02, ICC = 0.03, N (centreid) = 23, Observations = 13,458, Marginal R² = 0.087/ Conditional R² = 0.119,

**S10 Mixed-effects logistic regression examining the association between antimicrobial allergy labels and incidence of antimicrobial side effects within 30 days after surgery**

| **Characteristic**^1^ | **OR** **(95% CI)** | **p-value** |
| --- | --- | --- |
| **(Intercept)** | 0.01 (0.00 to 0.02) | **<0.001** |
| **Antimicrobial label** |  |  |
| No antimicrobial allergy label | — |  |
| Any antimicrobial allergy label | 0.93 (0.76 to 1.13) | 0.45 |
| **Age** |  |  |
| 18-37 | — |  |
| 38-57 | 1.19 (0.78 to 1.82) | 0.42 |
| 56-77 | 1.75 (1.14 to 2.67) | **0.010** |
| 78 and above | 2.83 (1.82 to 4.41) | **<0.001** |
| **Sex** |  |  |
| Female | — |  |
| Male | 1.38 (1.17 to 1.64) | **<0.001** |
| **Current Smoker** |  |  |
| Current Smoker(N) | — |  |
| Current Smoker(Y) | 0.95 (0.74 to 1.22) | 0.69 |
| **Surgical procedure** |  |  |
| Primary hip/knee replacement | — |  |
| Caesarean section | 0.72 (0.43 to 1.21) | 0.21 |
| Colorectal resection | 1.03 (0.77 to 1.37) | 0.85 |
| Hysterectomy | 0.62 (0.45 to 0.85) | **0.003** |
| Internal fixation of a closed long bone fracture | 1.08 (0.86 to 1.35) | 0.51 |
| Trans-urethral resection of prostate/bladder tumour | 0.58 (0.43 to 0.77) | **<0.001** |
| **Wound Contamination** |  |  |
| No | — |  |
| Yes | 1.60 (0.90 to 2.88) | 0.11 |
| **Surgery time (hrs)** |  |  |
| 0 - <2 hours | — |  |
| 2 - <4 hours | 1.45 (1.20 to 1.76) | **<0.001** |
| 4 - <6 hours | 1.93 (1.40 to 2.65) | **<0.001** |
| > 6 hours | 4.00 (2.84 to 5.63) | **<0.001** |
| **Frailty** |  |  |
| Non-frail | — |  |
| Mild Frailty | 1.34 (1.09 to 1.64) | **0.005** |
| Moderate Frailty | 1.63 (1.29 to 2.07) | **<0.001** |
| Severe Frailty | 2.36 (1.55 to 3.60) | **<0.001** |
| **ASA Physical Status Classification** |  |  |
| 1 | — |  |
| 2 | 1.77 (1.04 to 3.03) | **0.036** |
| 3 | 2.53 (1.46 to 4.40) | **<0.001** |
| 4 | 3.67 (1.96 to 6.87) | **<0.001** |
| **Diabetes Mellitus** | 1.46 (1.23 to 1.74) | **<0.001** |
| **Heart Failure** | 1.36 (1.04 to 1.78) | **0.027** |
| **Stroke** | 1.04 (0.77 to 1.39) | 0.80 |
| **Hypertension** | 1.37 (1.17 to 1.62) | **<0.001** |
| **Coronary Artery Disease** | 1.09 (0.88 to 1.35) | 0.44 |
| **Asthma/COPD** | 1.02 (0.84 to 1.23) | 0.87 |
| **Peripheral Vascular Disease** | 0.91 (0.61 to 1.35) | 0.64 |
| **Active Cancer** | 0.97 (0.80 to 1.18) | 0.78 |
| **Liver Cirrhosis** | 1.50 (0.73 to 3.07) | 0.27 |
| **Obesity** | 0.89 (0.73 to 1.08) | 0.24 |
| **centreid.sd__(Intercept)** | 0.42 (NA to NA) |  |

^1^Random Effects: σ² (Residual) = 3.29, τ00 (centreid) = 0.18, ICC = 0.05, N (centreid) = 23, Observations = 13,458, Marginal R²/Conditional R² = 0.228/0.267

Abbreviations: CI = Confidence Interval, OR = Odds Ratio

**S11 Mixed effects logistic regression examining the association between antimicrobial allergy labels and mortality within 30 days after surgery**

| **Characteristic**^1^ | **OR** **(95% CI)** | **p-value** |
| --- | --- | --- |
| **(Intercept)** | 0.00 (0.00 to 0.00) | **<0.001** |
| **Antimicrobial label** |  |  |
| No antimicrobial label | — |  |
| Any Antimicrobial | 0.95 (0.53 to 1.70) | 0.87 |
| **Sex** |  |  |
| Female | — |  |
| Male | 2.19 (1.38 to 3.46) | **<0.001** |
| **current smoker** |  |  |
| Current Smoker(N) | — |  |
| Current Smoker(Y) | 0.49 (0.19 to 1.22) | 0.12 |
| **wound contamination** |  |  |
| No | — |  |
| Yes | 1.45 (0.19 to 10.9) | 0.72 |
| **Frailty** |  |  |
| Non-frail | — |  |
| Mild Frailty | 3.56 (1.77 to 7.15) | **<0.001** |
| Moderate Frailty | 14.0 (7.66 to 25.6) | **<0.001** |
| Severe Frailty | 54.9 (26.9 to 112) | **<0.001** |
| **Diabetes Mellitus** | 0.87 (0.51 to 1.51) | 0.63 |
| **Heart Failure** | 0.70 (0.34 to 1.46) | 0.35 |
| **Stroke** | 0.89 (0.43 to 1.86) | 0.76 |
| **Hypertension** | 1.01 (0.64 to 1.59) | 0.97 |
| **Coronary Artery Disease** | 1.52 (0.88 to 2.60) | 0.13 |
| **Asthma/COPD** | 1.37 (0.82 to 2.26) | 0.23 |
| **Peripheral Vascular Disease** | 1.74 (0.79 to 3.84) | 0.17 |
| **Active Cancer** | 1.58 (0.99 to 2.53) | 0.055 |
| **Liver Cirrhosis** | 1.00 (0.13 to 8.03) | >0.99 |
| **Obesity** | 1.09 (0.60 to 1.97) | 0.77 |
| **centreid.sd__(Intercept)** | 0.35 (NA to NA) |  |
| **Characteristic**^1^ | **OR** **(95% CI)** | **p-value** |
| **(Intercept)** | 0.00 (0.00 to 0.00) | **<0.001** |
| **Antimicrobial label** |  |  |
| No antimicrobial label | — |  |
| Any Antimicrobial | 0.95 (0.53 to 1.70) | 0.87 |
| **Sex** |  |  |
| Female | — |  |
| Male | 2.19 (1.38 to 3.46) | **<0.001** |
| **current smoker** |  |  |
| Current Smoker(N) | — |  |
| ^1^Random Effects: σ² (Residual) = 3.29, τ00 (centreid) = 0.12, ICC = 0.04, N (centreid) = 23, Observations = 13,549, Marginal R²/Conditional R² = 0.259/0.286 | | |

**S12 Mixed-effects logistic regression examining the association between penicillin allergy labels and all postoperative complications within 30 days of surgery**

| **Characteristic**^1^ | **OR** **(95% CI)** | **p-value** |
| --- | --- | --- |
| **(Intercept)** | 0.05 (0.03 to 0.07) | **<0.001** |
| **Allergy_Categories** |  |  |
| No allergy | — |  |
| Non-penicillin | 1.19 (1.06 to 1.34) | **0.003** |
| Penicillin | 1.25 (1.09 to 1.43) | **0.002** |
| **Age** |  |  |
| 18-37 | — |  |
| 38-57 | 1.13 (0.94 to 1.35) | 0.20 |
| 56-77 | 1.26 (1.03 to 1.54) | **0.025** |
| 78 and above | 1.82 (1.45 to 2.27) | **<0.001** |
| **Sex** |  |  |
| Female | — |  |
| Male | 1.18 (1.05 to 1.33) | **0.006** |
| **current_smoker** |  |  |
| Current Smoker(N) | — |  |
| Current Smoker(Y) | 1.19 (1.03 to 1.37) | **0.017** |
| **Surgical procedure** |  |  |
| Primary hip/knee replacement | — |  |
| Caesarean section | 1.97 (1.55 to 2.50) | **<0.001** |
| Colorectal resection | 1.82 (1.51 to 2.20) | **<0.001** |
| Hysterectomy | 1.37 (1.14 to 1.64) | **<0.001** |
| Internal fixation of a closed long bone fracture | 1.01 (0.85 to 1.19) | 0.94 |
| Trans-urethral resection of prostate/bladder tumour | 1.11 (0.92 to 1.34) | 0.26 |
| **woundcontamination_operation** |  |  |
| No | — |  |
| Yes | 2.30 (1.54 to 3.43) | **<0.001** |
| **Surgery time (hrs)** |  |  |
| 0 - <2 hours | — |  |
| 2 - <4 hours | 1.40 (1.24 to 1.59) | **<0.001** |
| 4 - <6 hours | 1.75 (1.43 to 2.13) | **<0.001** |
| > 6 hours | 3.13 (2.48 to 3.95) | **<0.001** |
| **Frailty** |  |  |
| Non-frail | — |  |
| Mild Frailty | 1.14 (0.99 to 1.31) | 0.061 |
| Moderate Frailty | 1.40 (1.18 to 1.66) | **<0.001** |
| Severe Frailty | 3.29 (2.33 to 4.66) | **<0.001** |
| **ASA Physical Status Classification** |  |  |
| 1 | — |  |
| 2 | 1.31 (1.05 to 1.65) | **0.019** |
| 3 | 1.90 (1.49 to 2.43) | **<0.001** |
| 4 | 3.42 (2.41 to 4.86) | **<0.001** |
| **Diabetes Mellitus** | 1.24 (1.11 to 1.40) | **<0.001** |
| **Heart Failure** | 1.18 (0.95 to 1.47) | 0.14 |
| **Stroke** | 0.98 (0.79 to 1.23) | 0.88 |
| **Hypertension** | 1.08 (0.97 to 1.20) | 0.14 |
| **Coronary Artery Disease** | 1.05 (0.89 to 1.23) | 0.57 |
| **Asthma/COPD** | 1.19 (1.05 to 1.34) | **0.006** |
| **Peripheral Vascular Disease** | 0.98 (0.73 to 1.30) | 0.88 |
| ^1^Random Effects: σ² (Residual) = 3.29, τ00 (centreid) = 0.26 ICC = 0.07, N (centreid) = 23, Observations = 13,458, Marginal R² = 0.100/ Conditional R² = 0.165, | | |

**S13 Mixed-effects logistic regression examining the association between penicillin allergy labels and surgical site infections within 30 days of surgery**

| **Characteristic**^1^ | **OR** **(95% CI)** | **p-value** |
| --- | --- | --- |
| **(Intercept)** | 0.01 (0.01 to 0.02) | **<0.001** |
| **Allergy** |  |  |
| No allergy | — |  |
| Non-penicillin | 1.15 (0.97 to 1.38) | 0.11 |
| Penicillin | 1.25 (1.02 to 1.53) | **0.030** |
| **Age** |  |  |
| 18-37 | — |  |
| 38-57 | 0.86 (0.67 to 1.10) | 0.24 |
| 56-77 | 0.79 (0.60 to 1.04) | 0.10 |
| 78 and above | 0.78 (0.56 to 1.07) | 0.13 |
| **Sex** |  |  |
| Female | — |  |
| Male | 1.29 (1.07 to 1.56) | **0.007** |
| **Current smoker** |  |  |
| Current Smoker(N) | — |  |
| Current Smoker(Y) | 1.15 (0.94 to 1.41) | 0.17 |
| **Surgical procedure** |  |  |
| Primary hip/knee replacement | — |  |
| Caesarean section | 3.54 (2.42 to 5.18) | **<0.001** |
| Colorectal resection | 3.72 (2.72 to 5.08) | **<0.001** |
| Hysterectomy | 2.51 (1.84 to 3.43) | **<0.001** |
| Internal fixation of a closed long bone fracture | 0.87 (0.61 to 1.23) | 0.43 |
| Trans-urethral resection of prostate/bladder tumour | 6.26 (4.60 to 8.51) | **<0.001** |
| **Wound contamination** |  |  |
| No | — |  |
| Yes | 2.17 (1.33 to 3.55) | **0.002** |
| **Surgery time (hrs)** |  |  |
| 0 - <2 hours | — |  |
| 2 - <4 hours | 1.58 (1.26 to 1.96) | **<0.001** |
| 4 - <6 hours | 1.99 (1.47 to 2.68) | **<0.001** |
| > 6 hours | 2.98 (2.15 to 4.13) | **<0.001** |
| **Frailty** |  |  |
| Non-frail | — |  |
| Mild Frailty | 0.82 (0.66 to 1.02) | 0.069 |
| Moderate Frailty | 0.68 (0.50 to 0.93) | **0.016** |
| Severe Frailty | 1.45 (0.78 to 2.70) | 0.24 |
| **ASA Physical Status Classification** |  |  |
| 1 | — |  |
| 2 | 1.32 (0.95 to 1.84) | 0.10 |
| 3 | 1.89 (1.32 to 2.70) | **<0.001** |
| 4 | 2.73 (1.54 to 4.83) | **<0.001** |
| **Diabetes Mellitus** | 1.24 (1.05 to 1.47) | **0.012** |
| **Heart Failure** | 1.08 (0.75 to 1.57) | 0.67 |
| **Stroke** | 0.86 (0.59 to 1.25) | 0.43 |
| **Hypertension** | 0.89 (0.76 to 1.04) | 0.13 |
| **Coronary Artery Disease** | 0.99 (0.77 to 1.27) | 0.93 |
| **Asthma/COPD** | 1.06 (0.88 to 1.27) | 0.56 |
| **Peripheral Vascular Disease** | 0.70 (0.42 to 1.16) | 0.17 |
| **Active Cancer** | 0.84 (0.72 to 1.00) | **0.047** |
| **Liver Cirrhosis** | 1.00 (0.44 to 2.29) | >0.99 |
| **Obesity** | 1.44 (1.23 to 1.68) | **<0.001** |
| **centreid.sd__(Intercept)** | 0.54 (NA to NA) |  |

^1^Random Effects: σ² (Residual) = 3.29, τ00 (centreid) = 0.36 ICC = 0.10, N (centreid) = 23, Observations = 13,458, Marginal R² = 0.084/ Conditional R² = 0.174,

**S14 Mixed-effects negative binomial regression examining the association between penicillin allergy labels and all postoperative infections within 30 days after surgery**

| **Characteristic**^1^ | **OR** **(95% CI)** | **p-value** |
| --- | --- | --- |
| **(Intercept)** | 0.03 (0.02 to 0.05) | **<0.001** |
| **Allergy_Categories** |  |  |
| No allergy | — |  |
| Non-penicillin | 1.20 (1.06 to 1.37) | **0.005** |
| Penicillin | 1.30 (1.12 to 1.51) | **<0.001** |
| **Age** |  |  |
| 18-37 | — |  |
| 38-57 | 0.99 (0.81 to 1.20) | 0.92 |
| 56-77 | 1.12 (0.90 to 1.38) | 0.32 |
| 78 and above | 1.43 (1.12 to 1.82) | **0.004** |
| **Sex** |  |  |
| Female | — |  |
| Male | 1.15 (1.01 to 1.31) | **0.035** |
| **current_smoker** |  |  |
| Current Smoker(N) | — |  |
| Current Smoker(Y) | 1.31 (1.12 to 1.52) | **<0.001** |
| **Surgical procedure** |  |  |
| Primary hip/knee replacement | — |  |
| Caesarean section | 2.26 (1.74 to 2.94) | **<0.001** |
| Colorectal resection | 2.30 (1.87 to 2.83) | **<0.001** |
| Hysterectomy | 1.80 (1.48 to 2.20) | **<0.001** |
| Internal fixation of a closed long bone fracture | 1.01 (0.84 to 1.22) | 0.92 |
| Trans-urethral resection of prostate/bladder tumour | 1.47 (1.19 to 1.81) | **<0.001** |
| **woundcontamination_operation** |  |  |
| No | — |  |
| Yes | 2.09 (1.37 to 3.19) | **<0.001** |
| **Surgery time (hrs)** |  |  |
| 0 - <2 hours | — |  |
| 2 - <4 hours | 1.39 (1.20 to 1.60) | **<0.001** |
| 4 - <6 hours | 1.75 (1.42 to 2.17) | **<0.001** |
| > 6 hours | 2.63 (2.05 to 3.37) | **<0.001** |
| **Frailty** |  |  |
| Non-frail | — |  |
| Mild Frailty | 1.12 (0.96 to 1.31) | 0.13 |
| Moderate Frailty | 1.33 (1.10 to 1.60) | **0.004** |
| Severe Frailty | 2.96 (2.07 to 4.25) | **<0.001** |
| **ASA Physical Status Classification** |  |  |
| 1 | — |  |
| 2 | 1.25 (0.98 to 1.61) | 0.076 |
| 3 | 1.76 (1.34 to 2.31) | **<0.001** |
| 4 | 3.29 (2.26 to 4.79) | **<0.001** |
| **Diabetes Mellitus** | 1.15 (1.01 to 1.31) | **0.030** |
| **Heart Failure** | 1.03 (0.80 to 1.32) | 0.81 |
| **Stroke** | 1.06 (0.83 to 1.35) | 0.63 |
| **Hypertension** | 0.99 (0.89 to 1.11) | 0.90 |
| **Coronary Artery Disease** | 1.00 (0.84 to 1.20) | 0.97 |
| **Asthma/COPD** | 1.24 (1.09 to 1.41) | **0.001** |
| **Peripheral Vascular Disease** | 1.04 (0.76 to 1.42) | 0.82 |
| **Active Cancer** | 0.92 (0.81 to 1.05) | 0.22 |
| **Liver Cirrhosis** | 0.60 (0.30 to 1.22) | 0.16 |
| **Obesity** | 1.26 (1.12 to 1.42) | **<0.001** |
| **centreid.sd__(Intercept)** | 0.62 (NA to NA) |  |

^1^Random Effects: σ² (Residual) = 3.29, τ00 (centreid) = 0.39 ICC = 0.11, N (centreid) = 23, Observations = 13,458, Marginal R² = 0.083/ Conditional R² = 0.180,

**S15 Mixed-effects binomial regression examining the association between penicillin allergy labels and number of antimicrobial agents administered from recruitment until 30 days after surgery**

| **Group** | **Characteristic**^1^ | **exp(Beta)** **(95% CI)** | **p-value** |
| --- | --- | --- | --- |
| **cond** | (Intercept) | 1.35 (1.23 to 1.48) | **<0.001** |
|  | Allergy |  |  |
|  | Non-penicillin | 1.02 (0.99 to 1.06) | 0.22 |
|  | Penicillin | 1.16 (1.11 to 1.20) | **<0.001** |
|  | Age |  |  |
|  | Age38-57 | 0.97 (0.93 to 1.02) | 0.28 |
|  | Age56-77 | 1.00 (0.95 to 1.06) | 0.96 |
|  | Age78 and above | 1.07 (1.00 to 1.14) | **0.038** |
|  | Sex |  |  |
|  | Male | 1.04 (1.00 to 1.07) | **0.024** |
|  | Smoking status |  |  |
|  | Current Smoker(Y) | 1.03 (0.99 to 1.08) | 0.10 |
|  | Surgical procedure |  |  |
|  | Primary hip/knee replacement | — |  |
|  | Caesarean section | 1.26 (1.18 to 1.35) | **<0.001** |
|  | Colorectal resection | 1.51 (1.43 to 1.59) | **<0.001** |
|  | Hysterectomy | 1.12 (1.07 to 1.18) | **<0.001** |
|  | Internal fixation of a closed long bone fracture | 1.11 (1.06 to 1.17) | **<0.001** |
|  | Trans-urethral resection of prostate/bladder tumour | 0.88 (0.83 to 0.93) | **<0.001** |
|  | Wound contamination |  |  |
|  | wound contamination | 1.10 (0.98 to 1.23) | 0.10 |
|  | Surgery time (hrs) |  |  |
|  | 0 - <2 hours | — |  |
|  | 2 - <4 hours | 1.07 (1.03 to 1.10) | **<0.001** |
|  | 4 - <6 hours | 1.13 (1.07 to 1.19) | **<0.001** |
|  | > 6 hours | 1.35 (1.26 to 1.44) | **<0.001** |
|  | Frailty |  |  |
|  | Non-frail | — |  |
|  | Mild Frailty | 1.01 (0.97 to 1.05) | 0.69 |
|  | Moderate Frailty | 1.05 (1.00 to 1.11) | 0.051 |
|  | Severe Frailty | 1.23 (1.10 to 1.37) | **<0.001** |
|  | ASA Physical Status Classification |  |  |
|  | 1 | — |  |
|  | 2 | 1.07 (1.01 to 1.13) | **0.018** |
|  | 3 | 1.14 (1.07 to 1.21) | **<0.001** |
|  | 4 | 1.31 (1.18 to 1.44) | **<0.001** |
|  | Diabetes Mellitus | 1.03 (0.99 to 1.06) | 0.12 |
|  | Heart Failure | 1.00 (0.93 to 1.07) | 0.89 |
|  | Stroke | 1.03 (0.97 to 1.10) | 0.35 |
|  | Hypertension | 0.98 (0.95 to 1.01) | 0.29 |
|  | Coronary Artery Disease | 1.01 (0.96 to 1.06) | 0.71 |
|  | Asthma/COPD | 1.06 (1.02 to 1.09) | **0.003** |
|  | Peripheral Vascular Disease | 0.99 (0.91 to 1.08) | 0.84 |
|  | Active Cancer | 0.98 (0.95 to 1.02) | 0.27 |
|  | Liver Cirrhosis | 0.96 (0.81 to 1.14) | 0.63 |
|  | Obesity | 1.03 (1.0 to 1.06) | 0.10 |
|  | centreid.sd__(Intercept) | 0.12 (879,340 to 2,017,545,177) |  |

^1^Random Effects: σ² (Residual) = 0.43, τ00 (centreid) = 0.02, ICC = 0.03, N (centreid) = 23, Observations = 13,458, Marginal R² = 0.088/ Conditional R² = 0.119,

**S16 Mixed effects logistic regression examining the association between penicillin allergy labels and incidence of antimicrobial side effects within 30 days after surgery**

| **Characteristic**^1^ | **OR** **(95% CI)** | **p-value** |
| --- | --- | --- |
| **(Intercept)** | 0.01 (0.00 to 0.02) | **<0.001** |
| **Allergy Categories** |  |  |
| No allergy | — |  |
| Non-penicillin | 1.11 (0.93 to 1.32) | 0.27 |
| Penicillin | 0.88 (0.70 to 1.12) | 0.30 |
| **Age** |  |  |
| 18-37 | — |  |
| 38-57 | 1.19 (0.78 to 1.81) | 0.43 |
| 56-77 | 1.74 (1.14 to 2.66) | **0.010** |
| 78 and above | 2.81 (1.80 to 4.38) | **<0.001** |
| **Sex** |  |  |
| Female | — |  |
| Male | 1.39 (1.18 to 1.65) | **<0.001** |
| **current smoker** |  |  |
| Current Smoker(N) | — |  |
| Current Smoker(Y) | 0.96 (0.74 to 1.23) | 0.73 |
| **Surgical Procedure** |  |  |
| Primary hip/knee replacement | — |  |
| Caesarean section | 0.73 (0.44 to 1.23) | 0.24 |
| Colorectal resection | 1.03 (0.77 to 1.37) | 0.85 |
| Hysterectomy | 0.62 (0.45 to 0.85) | **0.003** |
| Internal fixation of a closed long bone fracture | 1.08 (0.86 to 1.36) | 0.48 |
| Trans-urethral resection of prostate/bladder tumour | 0.58 (0.44 to 0.78) | **<0.001** |
| **Wound contamination** |  |  |
| No | — |  |
| Yes | 1.61 (0.90 to 2.88) | 0.11 |
| **Surgery Time Category** |  |  |
| 0 - <2 hours | — |  |
| 2 - <4 hours | 1.45 (1.20 to 1.76) | **<0.001** |
| 4 - <6 hours | 1.93 (1.41 to 2.66) | **<0.001** |
| > 6 hours | 4.01 (2.85 to 5.65) | **<0.001** |
| **Frailty** |  |  |
| Non-frail | — |  |
| Mild Frailty | 1.34 (1.09 to 1.64) | **0.005** |
| Moderate Frailty | 1.63 (1.29 to 2.07) | **<0.001** |
| Severe Frailty | 2.36 (1.55 to 3.60) | **<0.001** |
| **ASA Score** |  |  |
| 1 | — |  |
| 2 | 1.77 (1.03 to 3.02) | **0.037** |
| 3 | 2.52 (1.45 to 4.38) | **0.001** |
| 4 | 3.65 (1.95 to 6.84) | **<0.001** |
| **Diabetes Mellitus** | 1.46 (1.22 to 1.73) | **<0.001** |
| **Heart Failure** | 1.36 (1.04 to 1.78) | **0.027** |
| **Stroke** | 1.04 (0.78 to 1.40) | 0.78 |
| **Hypertension** | 1.37 (1.17 to 1.61) | **<0.001** |
| **Coronary Artery Disease** | 1.08 (0.87 to 1.35) | 0.48 |
| **Asthma/COPD** | 1.01 (0.83 to 1.22) | 0.91 |
| **Peripheral Vascular Disease** | 0.91 (0.61 to 1.35) | 0.63 |
| **Active Cancer** | 0.97 (0.80 to 1.18) | 0.79 |
| **Liver Cirrhosis** | 1.48 (0.72 to 3.03) | 0.28 |
| **Obesity** | 0.89 (0.73 to 1.08) | 0.25 |
| **centreid.sd__(Intercept)** | 0.42 (NA to NA) |  |

^1^Random Effects: σ² (Residual) = 3.29, τ00 (centreid) = 0.18 ICC = 0.05, N (centreid) = 23, Observations = 13,458, Marginal R² = 0.228/ Conditional R² = 0.268,

Abbreviations: CI = Confidence Interval, OR = Odds Ratio
