## Supplementary material for "Drug allergy labels and complications after surgery: a prospective multi-centre cohort study": Investigator List

### Supplementary Files

**Investigator List**

**Writing Committee**

Tom E.F. Abbott, Louise Savic, Priyanthi Dias, Rupert M. Pearse, Salma Begum, Kamran Khan, Vikas Kaura

**Trial Management Group**

Louise Savic, Tom E.F. Abbott, Salma Begum, Priyanthi Dias, Kamran Khan, Vikas Kaura, Russell Hewson, Jai Vairale, Rupert M. Pearse

**Trial Steering Committee**

Tom E.F. Abbott, Louise Savic, Salma Begum, Priyanthi Dias, Alex Fowler, Vikas Kaura, Rupert. M. Pearse

**Local SAPPHIRE investigators:**

**Colchester Hospital, East Suffolk and North Essex NHS Foundation Trust**

Robert Lewis (Principal Investigator), Tracy Griffiths, Aashima Bakshi, Katrina Cooke, Chiara De Leonardis, Edyta Klata, Emma Williams, Kerry Smith, Michelle Dotchin

**Ipswich Hospital, East Suffolk and North Essex NHS Foundation Trust**

Robert Lewis (Principal Investigator), Vanessa Rivers, Caitlin Palmer

**East Cheshire NHS Foundation Trust**

Natalie Keenan (Principal Investigator), Christine Hibbert, Helen Wassall, Jan Tomkinson, Joanne Bradley-Potts, Katherine Rose, Philippa Hill, Sarah Ward, Hannah Brennan, Mohamed Mahmoud

**Pinderfields Hospital, Mid Yorkshire NHS Trust**

Brendan Sloan (Principal Investigator), Mark Southworth, Anna Littlejohns, Daniel Thomas,

Diana Kluczna, Nicholas Wroe, Tracy Langcake, James Roden, Chun Chiu, Jordon Almas,

Matthew Howe, Rachel Murali-Krishnan, Sarah Buckley, Elizabeth Denis, Thelma Darian, Alexandra Metcalfe, Amy Major, Abigail Crew

**North Bristol NHS Trust**

Ronelle Mouton (Principal Investigator), Amy Dodd, Alexander Ferriman, Carmel Oliver, Edward Mew, Kate Skidmore, Kat Herneman, Richard Elston, Stella Arthur Quarm, Timothy Walker

**Kingston Hospitals NHS Foundation Trust**

Gayani Jayasooriya (Principal Investigator), William Turner, Sejung Park, Isabel Bradley, Ramy Mahmoud, Sunitha Eswarappa, Roshni Molls, Amanda Davis, Judith Bradley, Margaret Grout, Jackie Latimer, Muqsith Rasool

**Barking, Havering and Redbridge University Hospitals NHS Trust**

Aparna George (Principal Investigator), George Joseph, Soumyajit Saha, Tatiana Pogreban, Toko Akangadjo, Mohammed Sabihul Islam, Sanjay K. Sakholiya, Amila Herath, Marvin Mutebi

**Royal United Hospitals, Bath NHS Foundation Trust**

Christopher Marsh (Principal Investigator), Alison Kirby, Andrew Ike, Annette Moreton, Catherine Bressington, Jenny Pullen, Jessica Sellick, Lowri Jones, Nora Seager-Wilkendorf,

Ros Knight, Rose Warren, Sara Burnard, Spandana Gumpula, Wendy Duberry

**The Royal Marsden NHS Foundation Trust**

Shaman Jhanji (Principal Investigator), Holly Townsend-Renn, Anna Warrington, David Parkinson, Ethel Black, Holly Hogan, Imran Ali, Maxime Caron-Goudreau

**Medway Maritime Hospital NHS Foundation Trust**

Kayleigh Jessop (Principal Investigator), Samantha Black, Dilukshi Wickramasinghe, Linda Ofori, Lisa Parker, Patience Nkala, Thomas Gower, Tom Hatton, Trudi Chronnell, Sam Martin, George Clews, Victoria Ewasiuk, Anais Mutarambirwa, Clarissa Madla

**The Rotherham NHS Foundation Trust**

Anil P. Hormis (Principal Investigator), Rachel Walker, Caroline Bird, Cheryl Graham, Laura Naish, Samuel Yale, Alasdair Sandland

**University Hospitals Birmingham NHS Foundation Trust**

Joyce Yeung (Principal Investigator), Helen Taylor, Jo Gresty, Juliet Sebastian, Mehwaish Zulfiqar, Meirvaan Basra, Fred Parker, Teresa Melody, Eleanor Reeves, Celina Maliaykal

**Croydon Health Services NHS Trust**

Sundar Ashok, Cassandra George, Christopher Black, Islam Abousharkh, Layla Guscoth, Louise Phaure, Sophie Boles, Sreedevi Rajalingam

**Newcastle upon Tyne Hospitals NHS Foundation Trust**

Rhona Sinclair (Principal Investigator), Iain J. McCullagh (Co-Principal Investigator), Rikzing D. Bhutia, Sabrina Kapur, Wendell Storr, Adam Cookson, Adam Mowatt, Alex Wilson, Arti Gulati, Anna McHugh, Anna Surridge, Annie Newby, Benjamin Brown, Benjamin Dowdell, Cecily Christopher, Chitra Garg, George Dykes, Hezekiah Awosusi, Ian J. Storey, Jake Taylor, James Newton, Jamie McPherson, Kirsty MacLeod, Laura Heggie, Maria Mazza, Martha Smith, Melissa Hartley, Nabeel Siddiqui, Natalie Hickling, Peter Bye

**Whipps Cross Hospital, Barts Health NHS Trust**

Tom EF. Abbott (Principal Investigator), Basil Nourallah (Co-Principal Investigator), Georgina Harridge (Co-Principal Investigator), Lucy Stephenson (Co-Principal Investigator), Jeveria Raja (Co-Principal Investigator), Laura Fulton, Juri Althonayan, Omar S. Ghori, Adele Mazzoleni, Aleksander Lysomirski, Fariha Rehman, Greg Titterton, James Tavner, Panayiotis Stavrinou, Ron Thomas, Salma Begum, Sarah-Louise Watson

**The Royal London Hospital, Barts Health NHS Trust**

Tom EF Abbott (Principal Investigator), Jonathan G Brend, (Co-Principal Investigator), Sukhmani Sra (Co-Principal Investigator), Imran Ali (Co-Principal Investigator), James Matthews (Co-Principal Investigator), James Waiting (Co-Principal Investigator), Michael Sawaryn, Adele Mazzoleni, Aleksander Lysomirski, Alexander Ka Jun Lee, Alexey Koroshilov, Dijay Dave, Evangelia Giannas, Fariha	Rehman, Fatima Seidu, Greg Titterton, James	Tavner, Juri Althonayan, Kavi Thobani, Ler Tu Seah, Mareena Joseph, Omar S. Ghori, Onika Ottley, Panayiotis Stavrinou, Rakshanda Mackay, Sarah-Louise Watson, Shreya Suresh, Sophie Arthur, Ron Thomas, Jai Vairale, Timothy Martin

**Newham University Hospital, Barts Health NHS Trust**

Bhavesh C. Gohil (Principal Investigator), Anjali Char (Co-Principal Investigator), Aderonke Adesanoye, Alice Gao, Eamon Dhall, Francis Elechi, Freddie Weyman, Deirdre Guerin, Harry Porter, James Lambert, Lynden Nicely, Michael Sawaryn, Ruth Parker

**St James's University Hospital, The Leeds Teaching Hospitals NHS Trust**

Louise Savic (Principal Investigator), Amy Longhurst, Anna Littlejohns, Beverley Jackson, Caroline Thomas, Catherine Moriarty, Chandan Gupta, Daniel Ruane, Jack Sherlock, Joe Gleeson-Buddhdev, Kapeel Dave, Lucy Marshall, Matthew Gowshall, Mohannad Mohyeldin, Nicholas Charlesworth, Nilmini Manawaduge, Samuel Flatau, Roshan S. Verghese

**Leeds General Infirmary, The Leeds Teaching Hospitals NHS Trust**

Vikas Kaura (Principal Investigator), Adam Neep, Anagha Joy, Ayman Salmona, Bridie O’neill, Chloe Uffendell, Dawne Conlon, Himani Pahuja, Holly Davies, Mark Priestley, Naina Stannard, Neha Hudlikar, Oliver Ross-Smith, Rachel Holmes, Rebecca King, Rosie Wragg

**Chapel Allerton Hospital/Wharfedale Hospital, The Leeds Teaching Hospitals NHS Trust**

Komal Ray (Principal Investigator), Amelia Milton, Aneesha Qadeer, Emma Walshaw, John Paul, John Wylde, Judith Sharp, Kanmani T. Lakshmikantha, Kate Gallagher, Michelle Naylor, Samuel Craven, Sneha Raju, Tetiana Khinalska, Zahid Furqan

**University Hospital Southampton NHS Foundation Trust**

Mark R. Edwards (Principal Investigator), Norma Diaper, Alice Baker, Karen Salmon, Sara Navalesi, Missy Harrison

**York and Scarborough Teaching Hospitals NHS Foundation Trust**

Katie Ayyash (Principal Investigator), Alison Cairns, Bryony Shelton, Hao Ming Kevin Cheung, Edward Kenny

**Chelsea and Westminster Hospital NHS Foundation Trust**

Ioannis Panagopoulos (Principal Investigator), Naoroz Patell, Pallavi Thakare, Thin Swe, Amrinder Sayan, Jamie Gonzales

**Royal Gwent Hospital, Aneurin Bevan University Health** **Board**

Tamas Szakmany (Principal Investigator), Aled Rees, Angharad King, Ashley Davies, Daniel Akintelure, Luke Agace, Mai Nur Sariah M. Nasser, Mary Perkins, Robert Yates

**Homerton University Hospital NHS Foundation Trust**

Chiraag Talati (Principal Investigator), Deborah Elf (Co-Principal Investigator), Alexander Rossides, (Co-Principal Investigator), Naomi Watson (Co-Principal Investigator), Debadutta Behera, Jumainah F. Rahman

**Walsall Manor Hospital NHS Trust**

Jacqueline Davies (Principal Investigator), Alex Bird, Jade Holmes, Joanne Arnold
